# Harnessing Machine Learning for Antimicrobial Resistance Surveillance in Zimbabwe

**DOI:** 10.1101/2025.09.02.25334969

**Authors:** Liberty Mutahwa, Hilary Takunda Takawira, Tinashe Muteveri, Perkins Watambwa, Delson Chikobvu, Whatmore Sengweni, Claris Siyamayambo, Justice Kasiroori, Lyson Chaka, Farai Mlambo

**Affiliations:** Midlands State University, Department of Applied Biosciences and Biotechnology, Gweru, Zimbabwe; University of Zimbabwe, Department of Mathematics and Computational Sciences, Harare, Zimbabwe; University of the Free State, Bloemfontein, South Africa; Midlands State University, Department of Applied Mathematics and Statistics, Gweru, Zimbabwe; South Africa Medical Research Council/ University of Johannesburg (SAMRC/UJ) Pan African Centre for Epidemics Research (PACER) Extramural Unit, Faculty of Health Sciences, South Africa; University of Zimbabwe, Department of Analytics and Informatics, Harare, Zimbabwe; University of Pretoria, Department of Statistics, Pretoria, South Africa; University of the Witwatersrand, Graduate School of Business Administration, Johannesburg, South Africa

**Keywords:** Antibiotic effectiveness, antimicrobial resistance, machine learning, multidrug resistance, support vector machine, surveillance, temporal trends

## Abstract

Antimicrobial resistance (AMR) poses a significant public health challenge, particularly in resource-limited settings such as Zimbabwe, where surveillance systems are often underdeveloped. This study aims to characterise AMR patterns at the Gweru provincial hospital (GPH) and evaluate machine learning (ML) models for predicting resistance to enhance surveillance. This retrospective cross-sectional study comprised 4 054 clinical isolates from 874 patient records (2022–2024). Five ML models, namely, support vector machine (SVM), random forest, logistic regression, gradient boosting, and k-nearest neighbors (KNN), were trained and evaluated, focusing on predictive performance for surveillance purposes. Among all evaluated models, SVM achieved the highest accuracy (72.08%), precision (73.25%), recall (79.78%), F1 score (0.76), and AUC-ROC (0.79), indicating it as the most effective model for AMR surveillance in this study. Feature importance analysis revealed that antibiotic class, hospital ward, patient age, and pathogen type were significant predictors of resistance. Notably, resistance was high for tetracycline (72.1%) and nitrofurantoin (75.7%), whereas imipenem (7.7%) showed the lowest resistance rates. Multidrug resistance was high among *S. aureus* (30%), whereas *Shigella* spp. and *Serratia marcescens* showed no multidrug resistance. This study highlights the significant AMR burden in Gweru and demonstrates the potential of ML, particularly SVM, for use in predictive surveillance. These findings support targeted interventions in high-risk hospital wards against specific pathogens, offering a scalable approach to AMR monitoring in resource-limited settings.

## 1. Introduction

Antimicrobial resistance has become one of the most significant global health issues of the 21st century, compromising the efficacy of treatments for infections that would otherwise be treatable (Bo *et al.,* 2024). Extensive overprescription and abuse of antibiotics in clinical practice have hastened the emergence of resistant bacterial populations. In 2019, AMR was directly responsible for an estimated 1.27 million fatalities globally (Aslam *et al*., 2024). If current trends continue, it is estimated that antimicrobial resistance can lead to as many as 10 million deaths each year by 2050, with Africa likely to face the highest burden, up to 4.5 million deaths each year (Sartorius *et al*., 2024). The projected economic burden of antimicrobial resistance is also significant, with an estimated worldwide cost of US$100 trillion (Murray *et al*., 2022). The burden will disproportionately be experienced in sub-Saharan Africa, where healthcare systems are already under pressure and infrastructure is lacking (Sartorius *et al*., 2024).

Across Africa, the AMR crisis has been driven by a mix of socioeconomic and systemic drivers. Pervasive poverty impacting adherence to expensive medications, frail health systems, and sometimes uncontrolled antibiotic dispensing has fostered conditions conducive to the rapid spread of resistance (Kariuki *et al*., 2022; Gulumbe *et al*., 2022). Africa has the world’s highest AMR-attributed death rate, 27.3 per 100 000 people, surpassing the combined fatalities of HIV/AIDS, tuberculosis, and malaria (Africa CDC, 2023). Despite this catastrophic burden, fewer than one-quarter of sub-Saharan African countries have rolling national action plans to combat AMR (Aruhomukama and Nakabuye, 2023). Zimbabwe is a typical example of these regional challenges. Mhondoro *et al*. (2019) reviewed more than 23 000 clinical bacterial isolates, identified *E. coli* (43.2%) and *S. aureus* (15.8%) as the most prevalent pathogens, and antibiotic resistance to the most widely used drugs like ampicillin and penicillin was in excess of 70%.

In Zimbabwe, a total of 3 900 deaths reported in 2021 were directly caused by AMR, and 15,800 deaths were attributed to resistant infections, which signified an increasing public health and economic burden (IHME, 2023). While in Africa, a few nations have made progress in digitised AMR surveillance, such as South Africa’s GERMS-SA, Kenya’s Fleming fund initiative (NICD, 2023; Chetty *et al*., 2022; Okolie *et al*., 2025). Zimbabwe has adopted a multisectoral One Health approach to promote collaboration in the human health, agriculture, and environmental sectors (Ren *et al*., 2022; Shapiro *et al*., 2019). However, its implementation has been slow and hindered by the lack of real-time digital surveillance systems (Pearcy *et al*., 2021). Zimbabwe’s AMR surveillance remains largely reactive, relying on traditional statistics, which primarily focus on severe cases like multidrug-resistant *K. pneumoniae* in intensive care units. These methods lack predictive capacity, hindering timely interventions (Kim *et al*., 2022). Such limitations are particularly evident in a medium-sized city such as Gweru, where health facilities often operate with minimal staff and inadequate resources for monitoring resistance patterns. The need for increased surveillance is indicated by Mhondoro *et al*. (2019), who reported elevated resistance levels in *S. aureus*, *E. coli* to commonly used antibiotics including amoxicillin. These results point to the necessity of introducing more comprehensive and prescriptive AMR surveillance systems that can guide empirical treatment guidelines and antimicrobial stewardship programs.

Conventional analysis methods, such as linear regression, are not robust enough to handle the complex and high-dimensional data produced by microbiology laboratories (Chiwodza *et al*., 2023). Emerging advances in machine learning offer a transformative approach to AMR surveillance, especially in settings with abundant data but limited resources. Machine learning algorithms have the potential to sift through large and diverse datasets, uncovering complex patterns, correlations, and trends that might be missed by traditional statistical methods (Broschat *et al*., 2024; Kim *et al*., 2022; Ren *et al*., 2022; Sakagianni *et al*., 2023). Machine learning has demonstrated its predictive capacity, such as predicting resistance profiles in *E. coli* based on genomic data (Pearcy *et al*., 2021). For instance, a multicountry study involving Uganda, Nigeria, and Tanzania used whole genome sequencing (WGS) data to validate ML models, achieving up to 92% accuracy in predicting *E. coli* resistance to ampicillin (Nsubuga *et al*., 2024). In Nigeria, random forest and LightGBM models were applied to predict resistance gene profiles with 81% accuracy (Adedeji *et al*., 2025). Babirye *et al*. (2024) demonstrated ML-based prediction of tuberculosis drug resistance using WGS data, achieving 85% accuracy in forecasting rifampicin resistance in Uganda. However, in Zimbabwe, the use of ML in public health surveillance is still in its early stages. Existing models often rely on global datasets that do not adequately account for local epidemiological, clinical, and environmental factors influencing AMR (Sunuwar and Azad, 2021).

This study aimed to explore the potential of ML in improving AMR surveillance and prediction by generating actionable insights from clinical data in a resource-limited country like Zimbabwe. Machine learning models can support infection control measures in Zimbabwe by detecting resistance trends and forecasting future trends, thereby providing evidence-based support for decisions and actions that enhance patient outcomes. The objectives of the study include (i) to develop a predictive ML model for AMR surveillance using clinical data collected from Gweru Provincial Hospital, (ii) to evaluate and select the best performing ML models, and (iii) to identify key pathogens, resistance drivers, and temporal trends that can guide focused interventions suited to the healthcare environment in Zimbabwe. This study sought to equip healthcare professionals with the knowledge and skills necessary to make informed decisions and allocate resources effectively to fight against AMR.

## 2. Materials and Methods

### 2.1 Study design

This study applied the retrospective cross-sectional design by analysing historically antimicrobial susceptibility tests (AST) results from bacterial cultures of 874 patients at Gweru Provincial Hospital (GPH) collected from 2022 to 2024.

### 2.2 Variables under consideration

The main outcome of this research was the AST result of bacterial isolates from clinical specimens. The results were used to classify bacterial isolates into susceptible, intermediate, or resistant categories to particular antibiotics according to standardised interpretive criteria, such as those of the Clinical and Laboratory Standards Institute (CLSI). The AST reports provided essential information to assess the trend and burden of AMR among frequently isolated pathogens. Multidrug resistance (MDR) in this study was defined as resistance to at least one agent in three or more antimicrobial categories (Sartorius *et al*., 2024; Kasew *et al*., 2021). Resistance was coded as a binary representing resistant (1) vs susceptible (0). This outcome served as a key measure for mapping the prevalence of drug resistance, informing empirical treatment guidelines, and examining temporal trends in resistance proportions across different seasons of the year.

### 2.3 Inclusion and exclusion criteria

All patients with laboratory confirmed bacterial infections were included, regardless of age, sex, or clinical condition, with no demographic restrictions applied. Patients negative for bacterial infections were excluded to focus on AMR patterns. The study analysis followed a multiphase approach (Figure 1): data collection, preprocessing, model development, and evaluation (Asadi *et al*., 2020).

**Figure 1:**
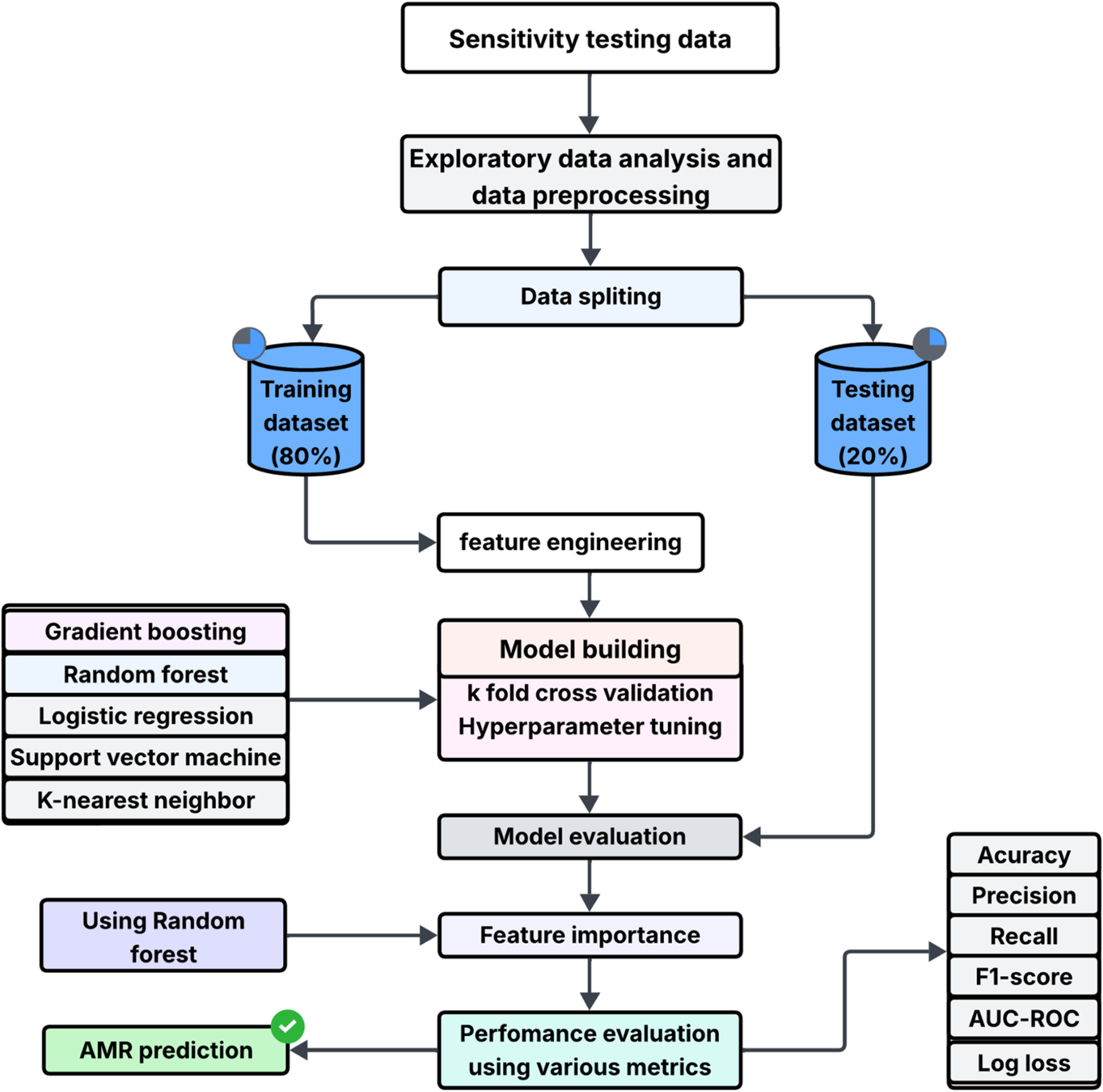
Flowchart illustrating the comprehensive approach for antimicrobial resistance prediction, detailing data processing, model development, and evaluation metrics.

### 2.4 Study setting

The study was conducted at Gweru Provincial Hospital, a general referral hospital in the Midlands Province, Zimbabwe. The GPH serves as a central hub for healthcare delivery, managing a high volume of infectious disease cases, including those requiring AST. The hospital’s diagnostic laboratory performs routine culture and susceptibility testing but faces challenges such as limited resources and a shortage of electronic data systems, which compromise AMR surveillance efforts. These conditions necessitated the development of a predictive model to enhance local monitoring capacity (Iskandar *et al*., 2021). Gweru Provincial Hospital serves both urban and rural populations, offering outpatient and inpatient services, as well as diagnostic microbiology. The hospital’s microbiology laboratory routinely processes specimens from various departments, including maternal, ICU, pediatric, and outpatient departments. The laboratory is equipped to perform standard culture and sensitivity testing, and it also receives referrals from various clinics across the Midlands province. Thus, GPH plays a critical role in regional surveillance of infectious diseases and antimicrobial resistance. These settings provided a valuable opportunity to examine real-world trends in bacterial infections and resistance patterns using routinely collected laboratory data.

### 2.5 Data source

All microbiological tests were conducted by the GPH laboratory as part of its diagnostic services; however, specimen collection, culture, identification, and antimicrobial testing were not performed in this study. Thus, this study was a secondary analysis of routine microbiological data generated at the GPH laboratory. Data were collected from laboratory worksheets with the results of AST for 874 patients. The datasets also contained demographic information and limited clinical histories (Table 1). Data was manually entered into a computerised database using LibreOffice Calc, and double-entry verification was performed to minimize transcription errors. Intermediate results (3%) were recorded as resistant (R) to ensure a binary outcome for model training, as this approach enhances model performance and helps to overcome class imbalance (Feretzakis *et al*., 2020).

**Table 1:** Overview of the independent variables in patient-related antimicrobial susceptibility tests.

| Variable | Description |
| --- | --- |
| Patient ID | A unique identifier for each patient used to track individual patients across multiple tests or records. It does not represent a unique sample, as one patient can have multiple rows. |
| Age group | The age category of the patient at the time of testing categorised into ranges (0-4, 5-17, 18-34, 35-49, 50-64, 65+). |
| Gender | The gender of the patient (male or female) |
| Ward | The hospital ward or clinic where the patient sample was collected (intensive care unit (ICU),outpatient department OPD). This indicates the location of care within the healthcare facility. |
| Clinical history | Medical history of the patient (Septic shock, urinary tract infection (UTI), No Clinical Records Provided (NCRP)). |
| Test date | The date when the AST was performed. |
| Seasons | Indicates the season during which the test was performed (summer or winter). |
| Specimen type | The type of clinical sample collected for testing (Blood, Urine, Pus, Swab, Cerebrospinal Fluid, HVS). |
| Pathogen name | The name of the microorganism identified (*S. aureus*, *E. coli*, *K. pneumoniae*, *C. albicans*). |
| Antibiotics/Drug | Antibiotic tested (Chloramphenicol, Vancomycin, Cotrimoxazole, Tetracycline). |

### 2.6 Data preprocessing

The initial dataset included 4 126 AST records (Figure 2). After excluding duplicate drug entries within individual sensitivity tests and removing rows with undefined drug codes, 4 054 isolates were retained for ML modeling and surveillance analysis.

**Figure 2:**
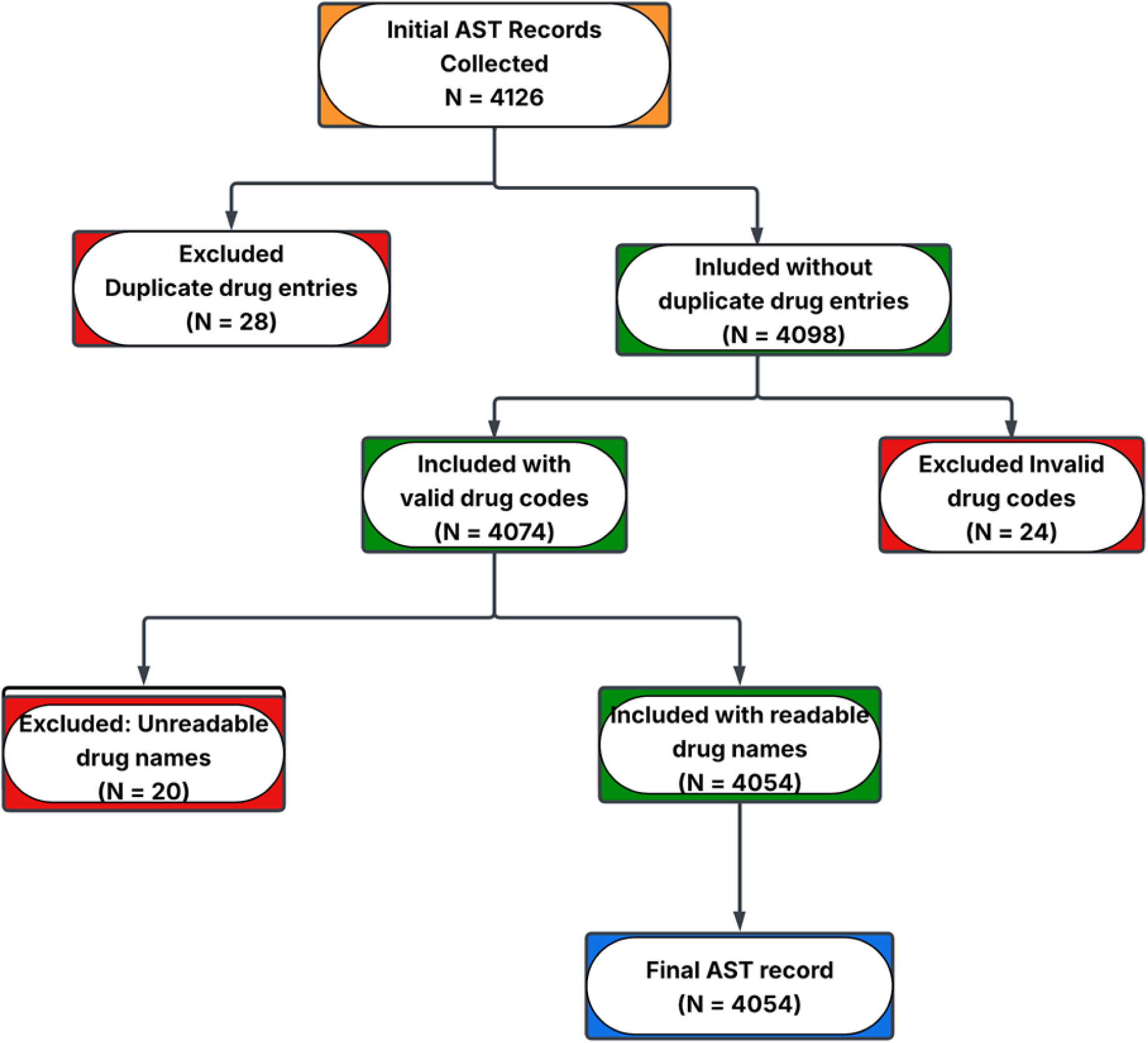
Flowchart illustrating the data cleaning process for antimicrobial sensitivity test records.

**Figure 3:**
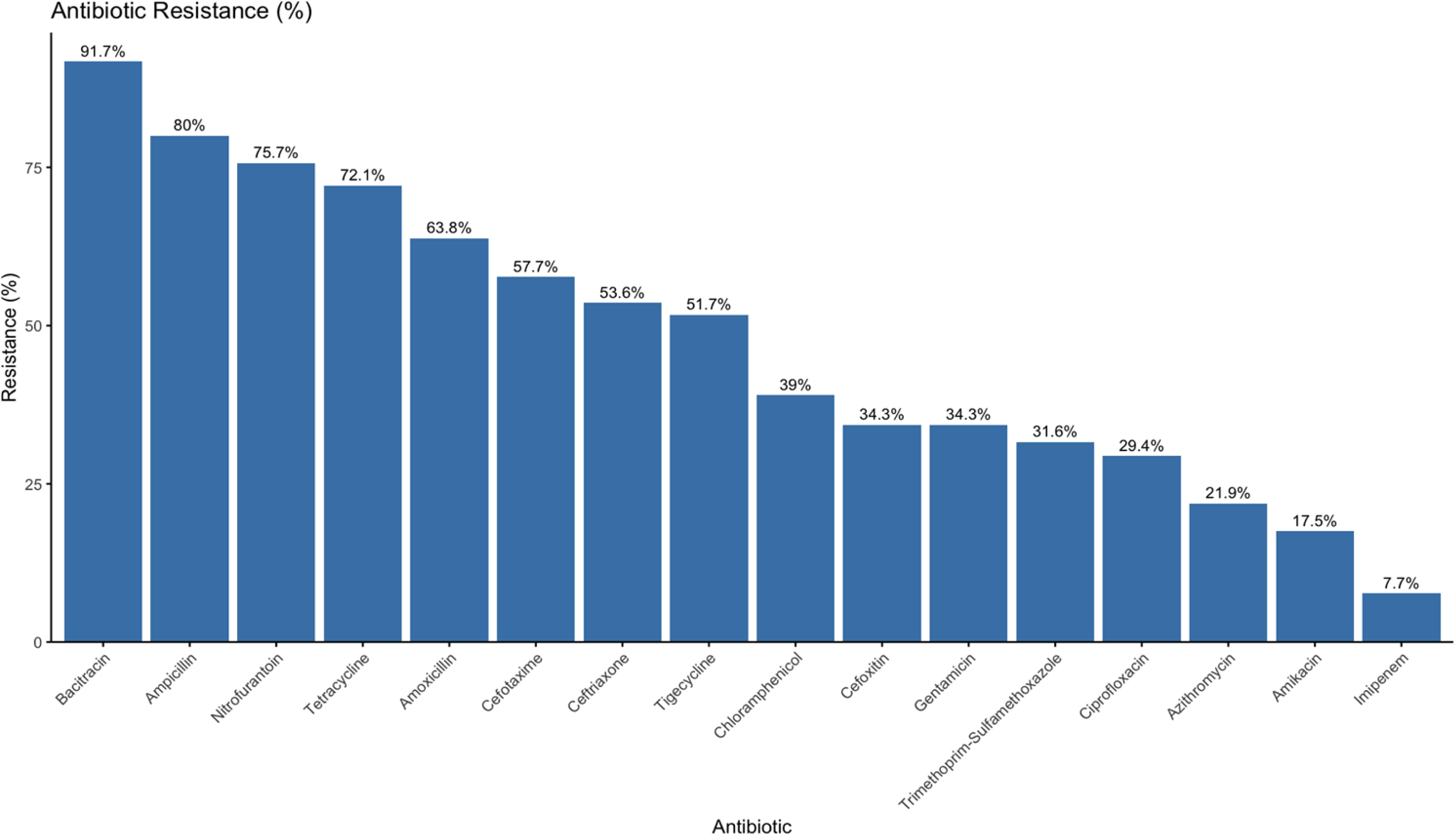
Percentage resistance of bacterial isolates to commonly tested antibiotics.

**Figure 4:**
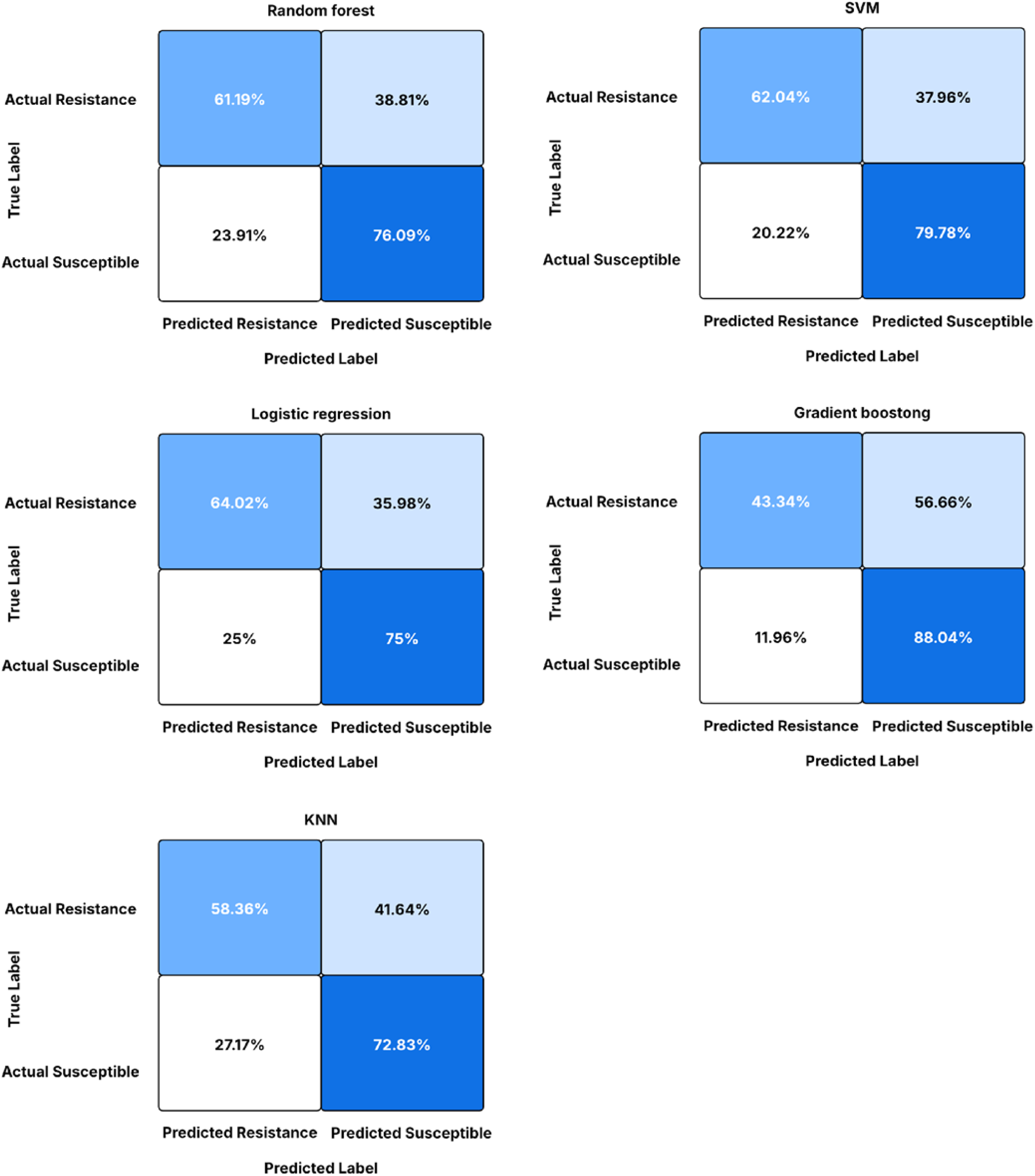
Confusion matrices of the performance of machine learning models.

Data preprocessing involved several steps to ensure compatibility with ML algorithms. The data format displayed variations, including inconsistent date formats, mixed string cases in categorical variables (for example, “Male” vs. “male”), and inconsistent naming of antibiotics and pathogens (abbreviations vs. full names). Continuous variables, including age, were categorised into age groups to capture non-linear relationships with resistance, which helped the model treat each age group as an independent entity. All categorical variables were transformed using one-hot encoding to convert non-numeric data into a format that models can process without incorrectly assuming an ordinal relationship between categories (Ali *et al*., 2023). The final dataset of 4 054 isolates was split into training (3 243 isolates; 80%) and testing (811 isolates; 20%) subsets. The split was performed using the train-test-split function from Scikit-learn (version 1.2.2) with a random seed (42) to ensure reproducibility.

### 2.7 Model building

Algorithm selection was guided by the nature of the dataset, interpretability of the models, and their capacity to perform AMR prediction tasks (Camacho *et al*., 2018). Logistic regression, SVM, KNN, and random forests were primarily considered because of their established effectiveness in classification (Khalid *et al*., 2023). Gradient boosting was also explored because it is known to enhance predictive accuracy and robustness by aggregating the predictions of multiple base models (Ganie *et al*., 2024). Five ML models (logistic regression, random forest, KNN, SVM and gradient boosting) were developed to predict AMR patterns from a cleaned dataset of 3 243 clinical isolate records from 874 patient records at GPH. All models were implemented in Python 3.10 using Scikit-learn (version 1.2.2) and pandas. A grid search combined with 10-fold stratified cross-validation was used to optimize model performance (Kasew *et al*., 2021). This approach tested different parameter combinations while preserving the resistant/susceptible class distribution across folds, ensuring fair and unbiased evaluation (Sartorius *et al*., 2024). The models, hyperparameters tuned, tested ranges, and rationale are shown in Table 2. Random forest was used to assess the impact of each feature on the final prediction of the ML model. This allowed the contribution of each variable to the model’s output to be determined (Quoc *et al*., 2023).

**Table 2:** Models selected and hyperparameter tuning.

| Model | Hyperparameter tuned | Values tested | Rationale |
| --- | --- | --- | --- |
| Logistic regression | C (Regularization strength) | 0.01, 0.1, 1, 10, 100 | Applied L2 regularization to reduce overfitting by penalizing overly large coefficients. |
| SVM | C (Regularization strength), $\gamma$ (Kernel coefficient) | C: 0.1, 1, 10 $\gamma$: 0.001, 0.01, 0.1 | Used an RBF kernel to capture non-linear patterns in the data. |
| Random forest | n_estimators (number of trees), max_depth (maximum tree depth) | n_estimators: (50, 100, 200),<br>max_depth: (10, 20, None) | Combined multiple decision trees to improve predictive stability and reduce overfitting. |
| Gradient boosting | learning_rate, n_estimators | learning_rate: 0.01, 0.1, 0.3<br>n_estimators: 50, 100 | Sequentially built models to correct prior errors for stronger predictions. |
| KNN | k (Number of neighbors) | 3, 5, 7, 9 | Classified isolates based on the most common label among nearest neighbors. |

### 2.8 Model evaluation

The training set (3 243 records) was partitioned into 10 equal folds, with each fold serving as a validation set once, whereas the remaining nine folds were used for training. Stratification preserved the balance between resistant and susceptible classes to address potential imbalances. Performance was evaluated on both the cross-validated training set and the hold-out test set (811 records) using a suite of metrics derived from confusion matrices (Table 3).

**Table 3:** Evaluation metrics based on positive and negative predictions of a model.

| Evaluation metric | Equation |
| --- | --- |
| Accuracy | $\frac{TP + TN}{TP + TN + FP + FN}$ |
| Specificity | $\frac{TN}{TN + FP}$ |
| Recall | $\frac{TP}{TP + FN}$ |
| F1-Score | $\frac{2 \times TP}{2 \times TP + FP + FN}$ |
| Log loss | $-\frac{1}{N} \sum_{i=1}^N [y_i \ln(p_i) + (1-y_i) \ln(1-p_i)]$ |

Where TP (true positive) was the number of resistant strains predicted to be resistant, TN (true negative) was the number of sensitive strains predicted to be sensitive, FP (false positive) was the number of sensitive strains predicted to be resistant, and FN (false negative) was the number of resistant strains predicted to be sensitive.

## 3. Results

### 3.1 Distribution of specimens and bacterial isolates

A total of 4 054 clinical specimens were examined to establish the distribution of specimen types and isolated pathogens. The most common specimens examined were pus samples (43.76%), followed by urine samples (32.04%), stool samples (7.89%), and blood samples (6.19%). The other specimens included swabs (3.43%), catheters (2.69%), high vaginal swabs (2.34%), pleural fluid (1.09%), cerebrospinal fluid (0.39%), septic wound swabs (0.12%), and ascitic fluid (0.05%) samples (Table 4).

**Table 4:** Frequency of specimen types and pathogens.

| Specimen information |  |  | Pathogen information |  |  |
| --- | --- | --- | --- | --- | --- |
| Specimen type | N | % | Pathogen name | N | % |
| Ascitic Fluid | 2 | 0.05 | *C. albicans* | 4 | 0.10 |
| Blood | 251 | 6.19 | *C. freundii* | 35 | 0.86 |
| CSF | 16 | 0.39 | *Clostridium spp.* | 7 | 0.17 |
| Catheter | 109 | 2.69 | *Corynebacterium spp.* | 3 | 0.07 |
| HVS | 95 | 2.34 | *Enterobacter spp.* | 41 | 1.01 |
| Pleural Fluid | 44 | 1.09 | *E. coli* | 358 | 8.83 |
| Pus (All) | 1 774 | 43.76 | *Influenzae ssp.* | 9 | 0.22 |
| Septic Wound | 5 | 0.12 | *K. pneumoniae* | 446 | 11.00 |
| Stool | 320 | 7.89 | Lactose Fermenting Coliforms | 1 068 | 26.34 |
| Swabs (All) | 139 | 3.43 | Non-Lactose Fermenting Coliforms | 338 | 8.34 |
| Urine | 1 299 | 32.04 | *P. mirabilis* | 175 | 4.32 |
|  |  |  | *Proteus vulgaris* | 28 | 0.69 |
|  |  |  | *Providencia spp.* | 4 | 0.10 |
|  |  |  | *P. aeruginosa* | 194 | 4.79 |
|  |  |  | *Salmonella typhi* | 16 | 0.39 |
|  |  |  | *Serratia marcescens* | 6 | 0.15 |
|  |  |  | *Shigella spp.* | 7 | 0.17 |
|  |  |  | *S. aureus* | 919 | 22.67 |
|  |  |  | *Streptococcus spp.* | 143 | 3.53 |
|  |  |  | *V. cholerae* | 253 | 6.24 |
| Total | 4 054 | 100.00 | Total | 4 054 | 100.00 |

The most common isolated pathogens were lactose-fermenting coliforms (LFC) (26.34%), *S. aureus* (22.67%), *K. pneumoniae* (11.00%), and *E. coli* (8.83%) (Table 4). *V. cholarae* represented 6.24% of isolates, reflecting potential cholera outbreaks, while the occurrence of non-LFCs (8.34%) and *P. aeruginosa* (4.79%) reflected the presence of opportunistic and potentially drug-resistant pathogens. Other less commonly isolated microorganisms included *P. mirabilis* (4.32%), *Streptococcus* spp. (3.53%), *C. freundii* (0.86%), *Enterobacter* spp. (1.01%), and *C. albicans* (0.10%).

Analysis of 4 054 AST results from 874 patients at GPH revealed varied resistance patterns across gender, age group, season, and antibiotic type. High resistance was observed in 45.5% of the isolates from males and 41.0% of the isolates from women. For the age group, resistance ranged from 35.1% in children aged 0–4 years to 45.4% in adults aged 65 years and older. Notable differences in resistance were recorded between seasons, with resistance occurring in 45.4% of the isolates in winter and 41.6% of the isolates in summer (Table 5).

**Table 5:**
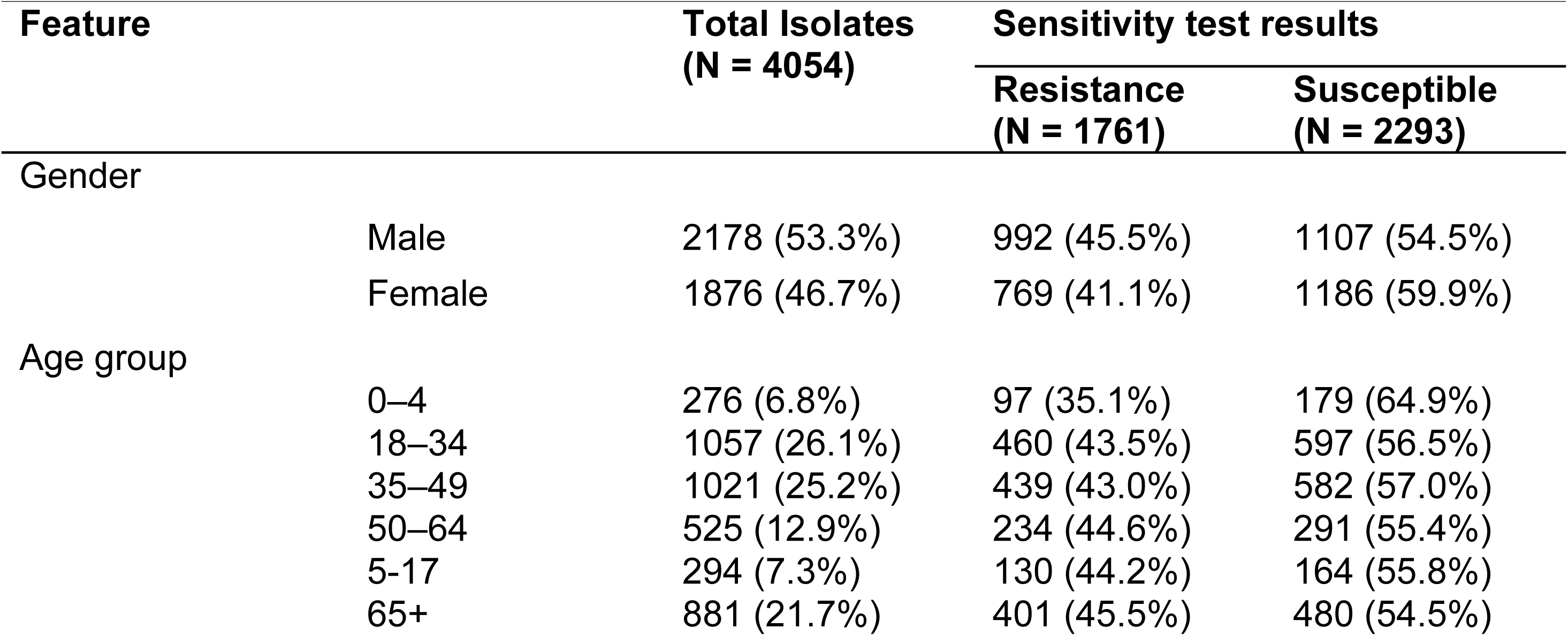
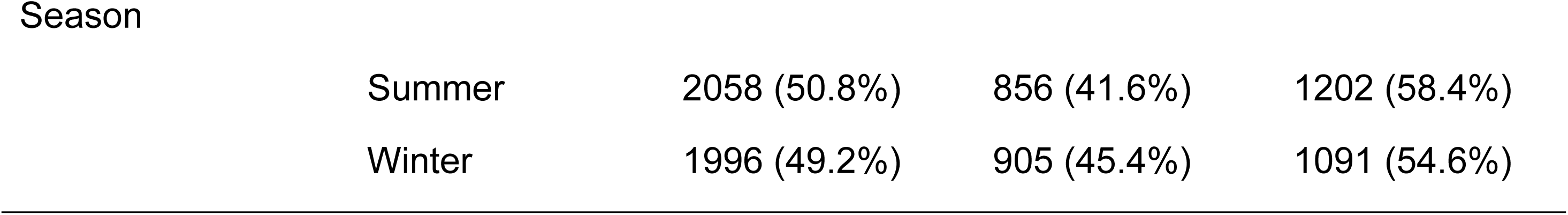
A summary of the distribution of resistance and susceptibility among various demographics and antibiotic treatments.

Resistance varied across antibiotics, with bacitracin (91.7%) being the highest, followed by ampicillin (80%) and nitrofurantoin (75.7%). Moderate resistance was observed for tetracycline (72.1%), amoxicillin (63.8%), and cefotaxime (57.7%). Low resistance levels were recorded for antibiotics such as amikacin (17.5%) and imipenem (7.7%). The trend highlights widespread resistance to older, commonly used antibiotics, while last-resort drugs retain greater efficacy.

### 3.2 Performance and evaluation of ML models

The ML models evaluated on the test set of 811 isolates (20%) demonstrated effectiveness in predicting resistance patterns for surveillance purposes. The SVM achieved the highest accuracy (72.1%), F1 score (0.764), and AUC-ROC (0.791), followed closely by logistic regression, which achieved a precision of 0.731 and log loss of 0.548. Gradient boosting excelled in recall (0.880), but had the lowest specificity (0.433), whereas random forest and KNN recorded balanced metrics (Table 6).

**Table 6:** Comparative performance metrics of random forest, logistic regression, SVM, gradient boosting, and KNN models for predicting antimicrobial resistance.

| Performance metrics | Random forest | Logistic regression | SVM | Gradient boosting | KNN |
| --- | --- | --- | --- | --- | --- |
| Accuracy | 0.6962 | 0.7023 | **0.7208** | 0.6863 | 0.6654 |
| Precision | 0.7187 | 0.7309 | **0.7325** | 0.6694 | 0.6950 |
| Recall | 0.7609 | 0.7500 | 0.7978 | **0.8804** | 0.7283 |
| F1 Score | 0.7392 | 0.7403 | **0.7638** | 0.7606 | 0.7113 |
| AUC-ROC | 0.7866 | 0.7890 | **0.7908** | 0.7767 | 0.7270 |
| Log Loss | 0.5530 | 0.5481 | 0.5463 | 0.6086 | **0.8908** |
| Specificity | 0.6119 | **0.6402** | 0.6204 | 0.4334 | 0.5836 |

Confusion matrices for the five ML models (random forest, logistic regression, SVM, Gradient boosting, and KNN) provided further insight into the classification performance for the resistance and susceptible classes. The SVM demonstrated the strongest balance, with a false negative rate of 20.22% (1 – recall), and a false positive rate of 37.96% (1 – specificity). Logistic regression achieved a moderate performance with a false positive rate of 25.00% and a false negative rate of 35.98%. Gradient boosting exhibited the highest recall (88%) and had a notably high false positive rate of 56.66% due to its low specificity. In contrast, random forest and KNN showed false positive rates of 23.91% and 27.17%, respectively, and false negative rates of 38.81% and 41.64% (Figure 4).

The ROC curve compares the classification performance of five models. SVM achieved the highest AUC (0.7886), followed closely by Logistic regression (0.7877), whereas KNN had the lowest (0.7532) (Figure 5).

**Figure 5:**
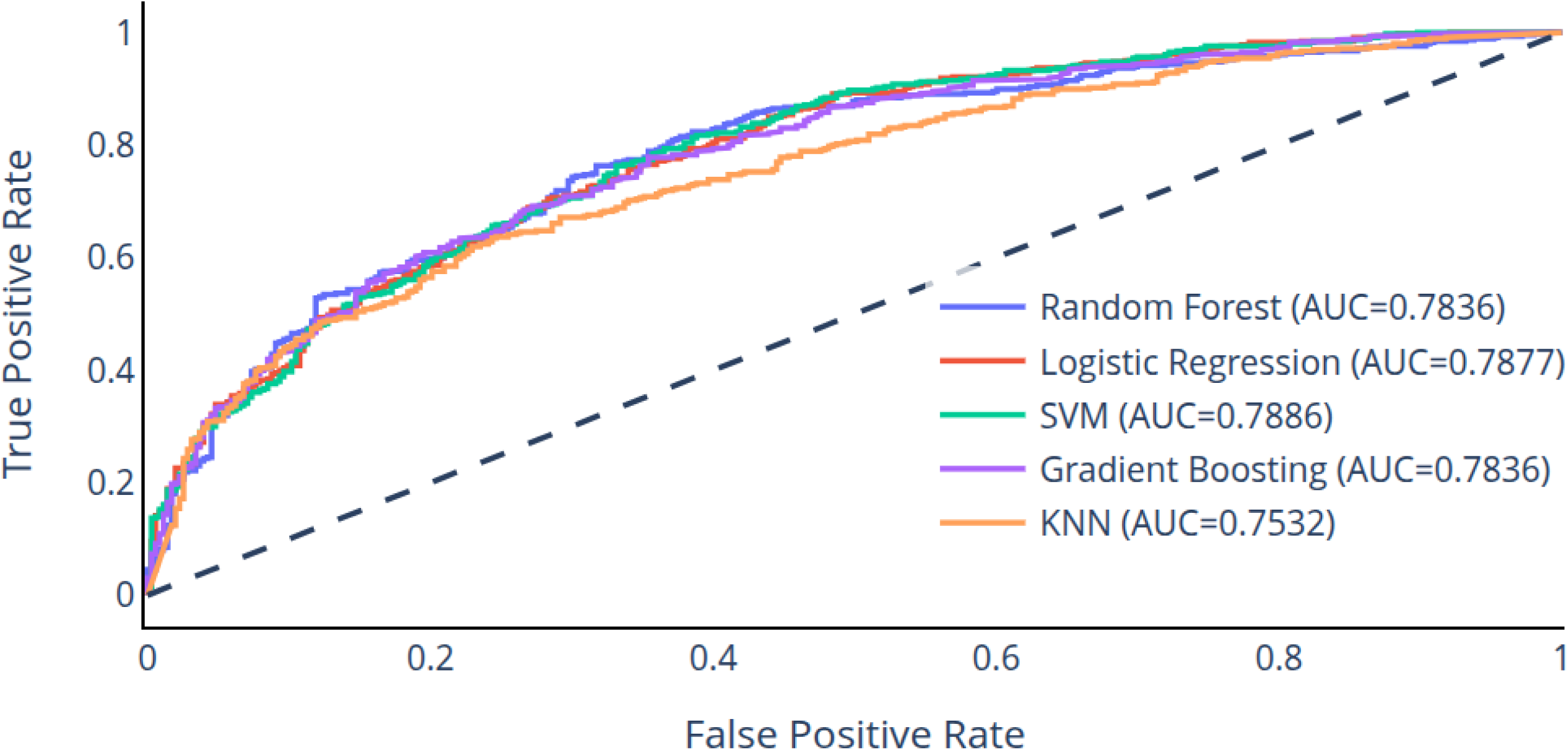
ROC-AUC curves for five machine learning models.

### 3.3 Analysis of key predictors and drivers of antimicrobial resistance

Feature importance analysis for the random forest model revealed that antibiotic type was the most influential predictor of resistance patterns, followed by the bacterial isolate type, with detailed importance scores ordered from highest to lowest (Figure 6).

**Figure 6:**
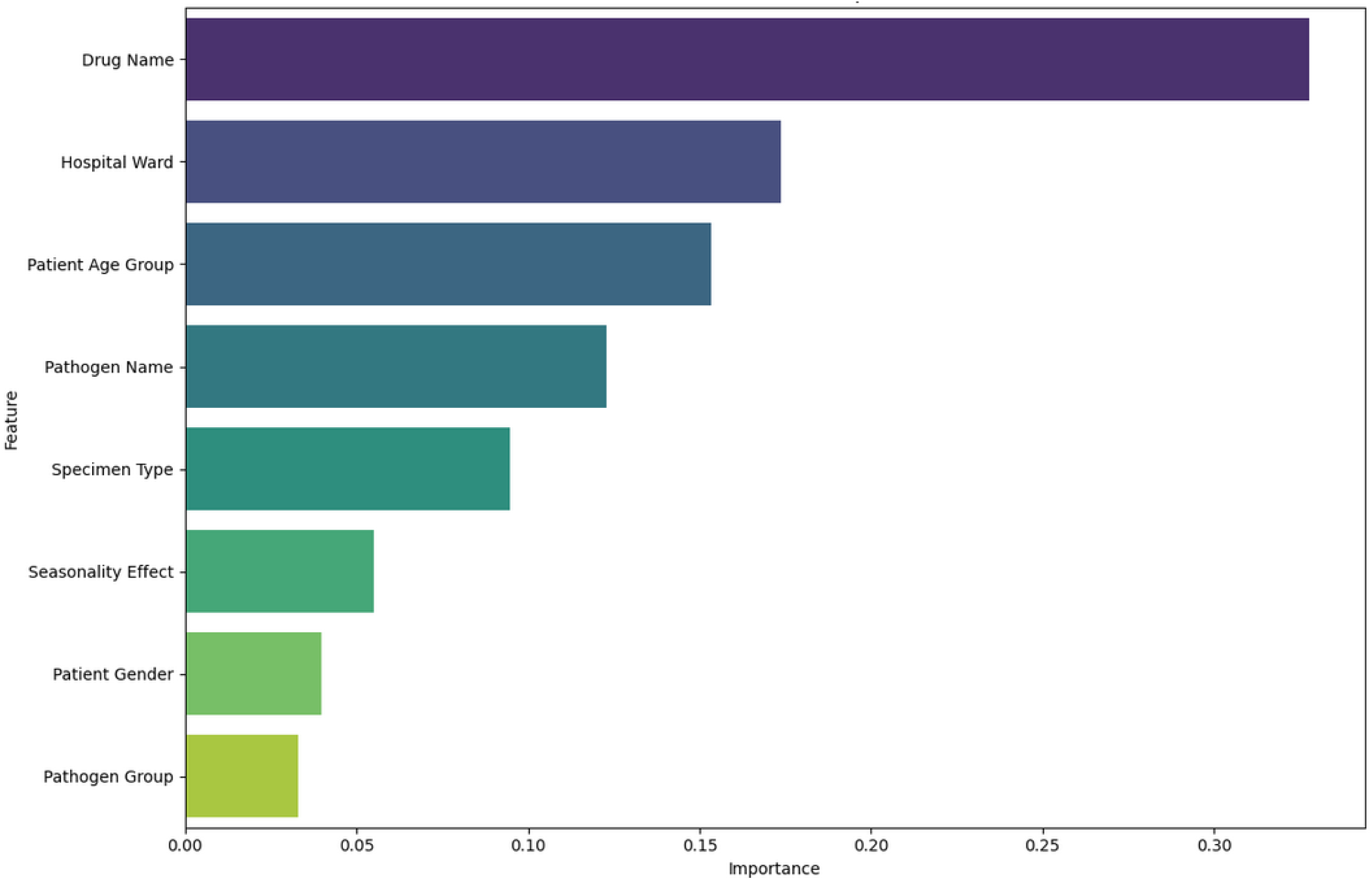
Feature importance from the random forest model.

Resistance rates differed significantly across hospital wards (Figure 7). The stroke ward had the highest median resistance rate of about 75%, whereas the postnatal ward had the lowest resistance rate of about 25%. Wards such as ICU and OPD displayed high variability, with resistance rates reaching 100% in some cases, highlighting areas of concern for targeted surveillance.

**Figure 7:**
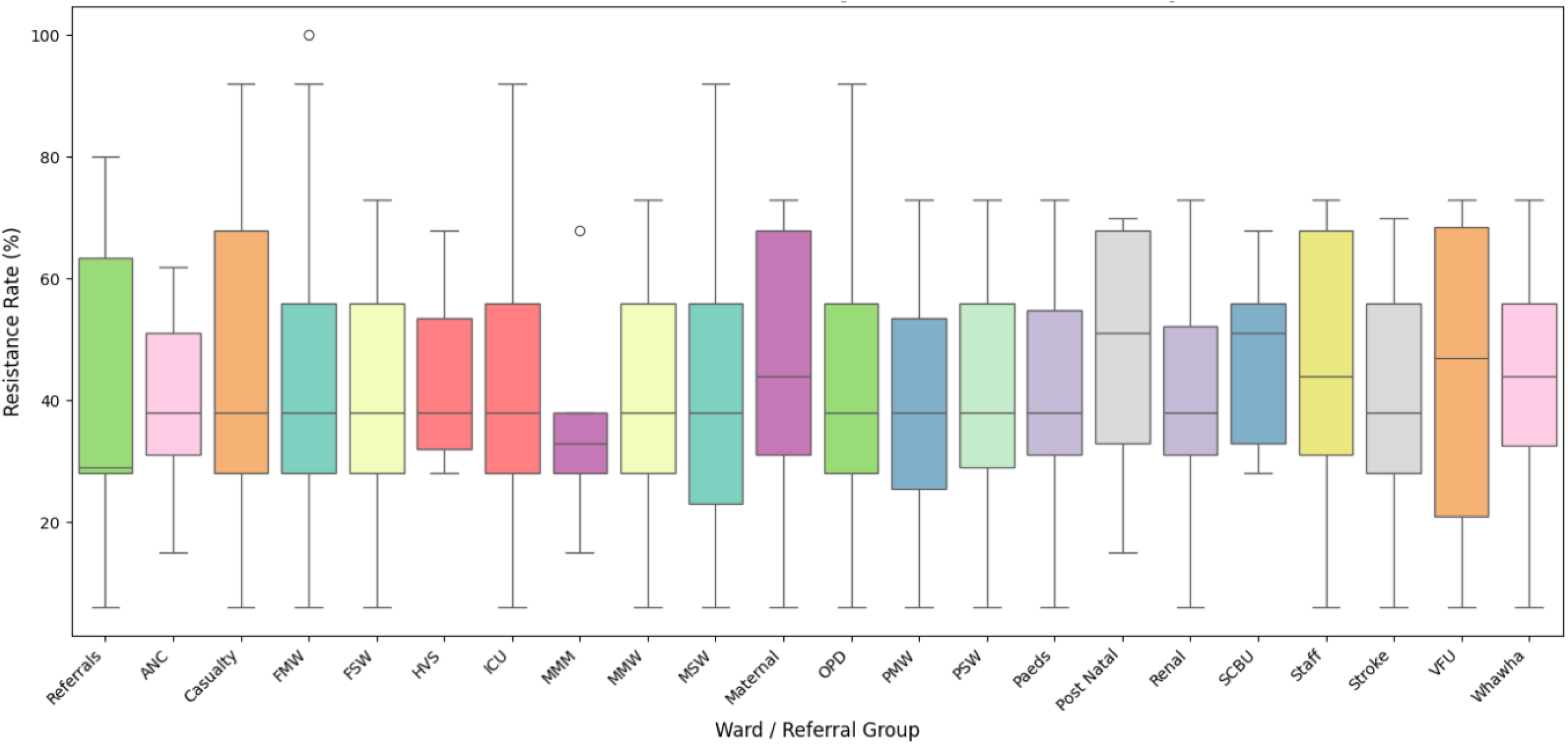
Resistance rates (%) of pathogens against antimicrobials across 21 wards and staff.

The resistance rates (%) of various pathogen classes to a specific antimicrobial agent are shown in Figure 8. *Providencia* spp. exhibited the highest resistance rate, reaching approximately 70%, while *Corynebacterium* spp. showed the lowest resistance at about 20%.

**Figure 8:**
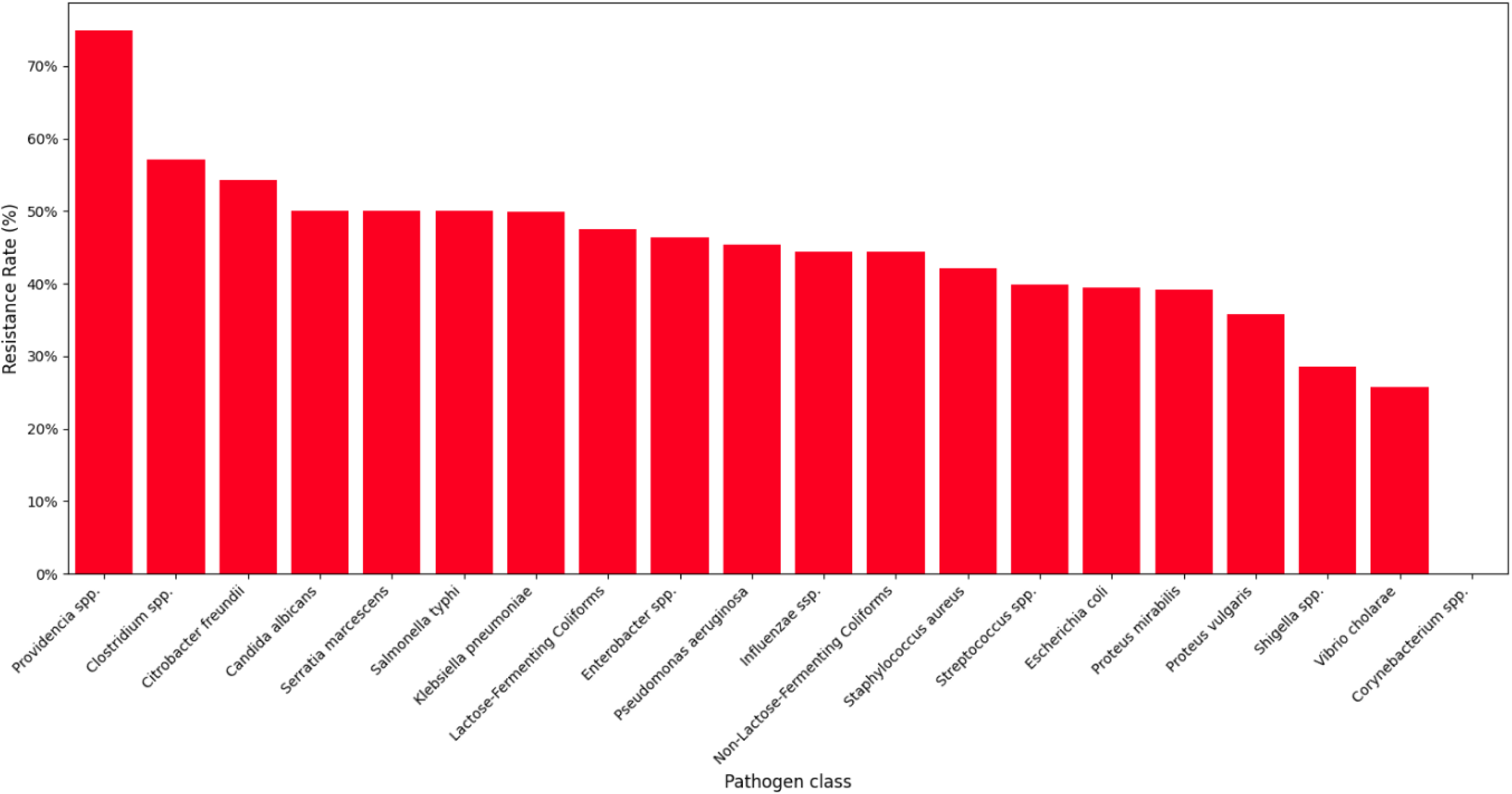
Resistance rates (%) of various pathogens to antimicrobial agents.

Antibiotic effectiveness against bacterial isolates, assessed using a 50% vulnerability threshold, varied across (Figure 9). Imipenem demonstrated the highest effectiveness at about 90%, while Bactrim was the least effective at about 10%. Most other drugs, including Ceftriaxone and Tetracycline, fell below the effectiveness threshold, indicating widespread AMR.

**Figure 9:**
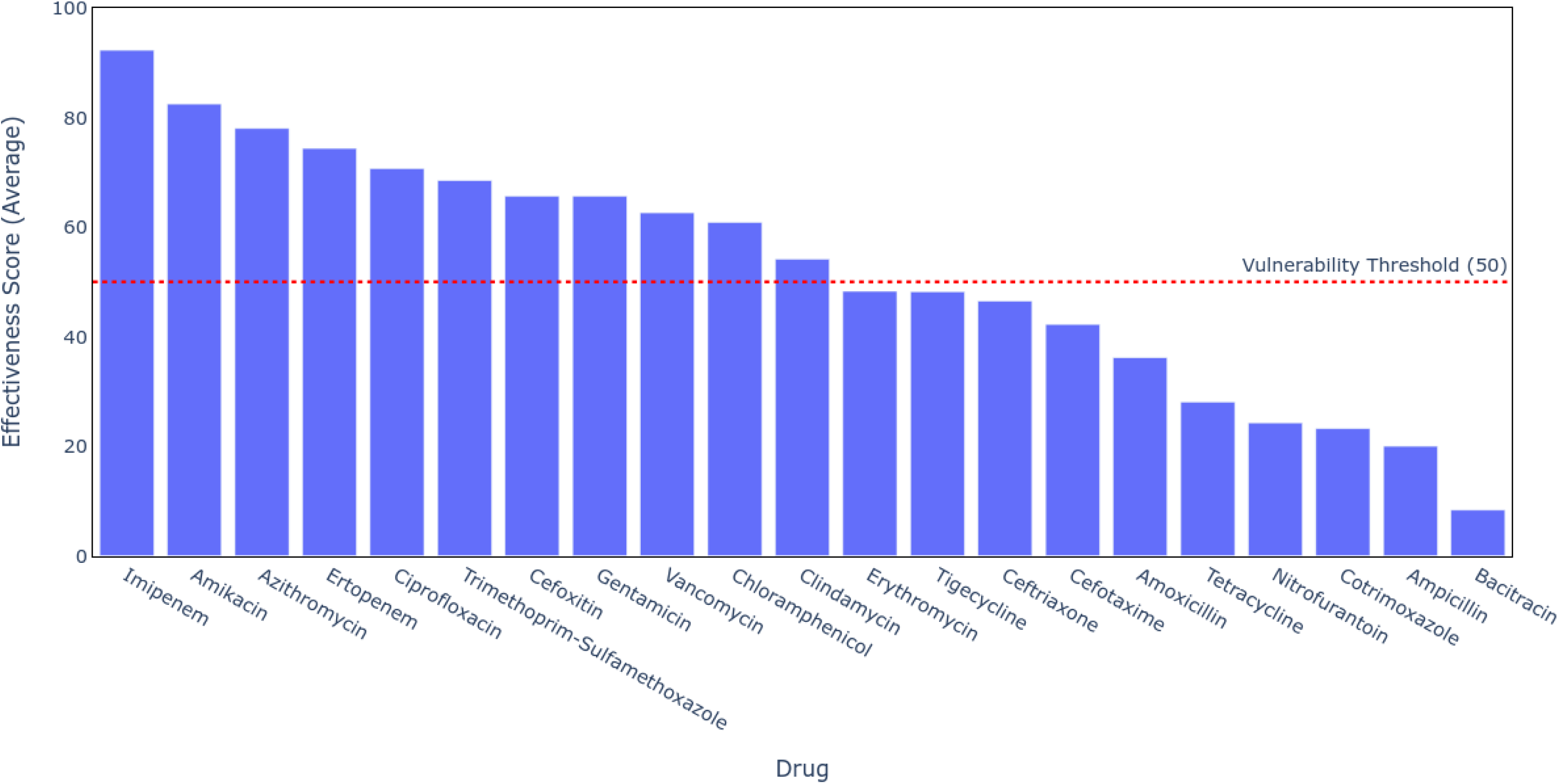
Effectiveness of antibiotics against isolated pathogens based on sensitivity tests and a vulnerability threshold of 50%.

Resistance patterns across bacterial isolates and antibiotics (Figure 10) underscore high resistance of *P. mirabilis* to most antibiotics (100% to amoxicillin and Ceftriaxone), in contrast to low resistance of *C. albicans* (0% to chloramphenicol). Specific rates are detailed in the heatmap (Figure 10).

**Figure 10:**
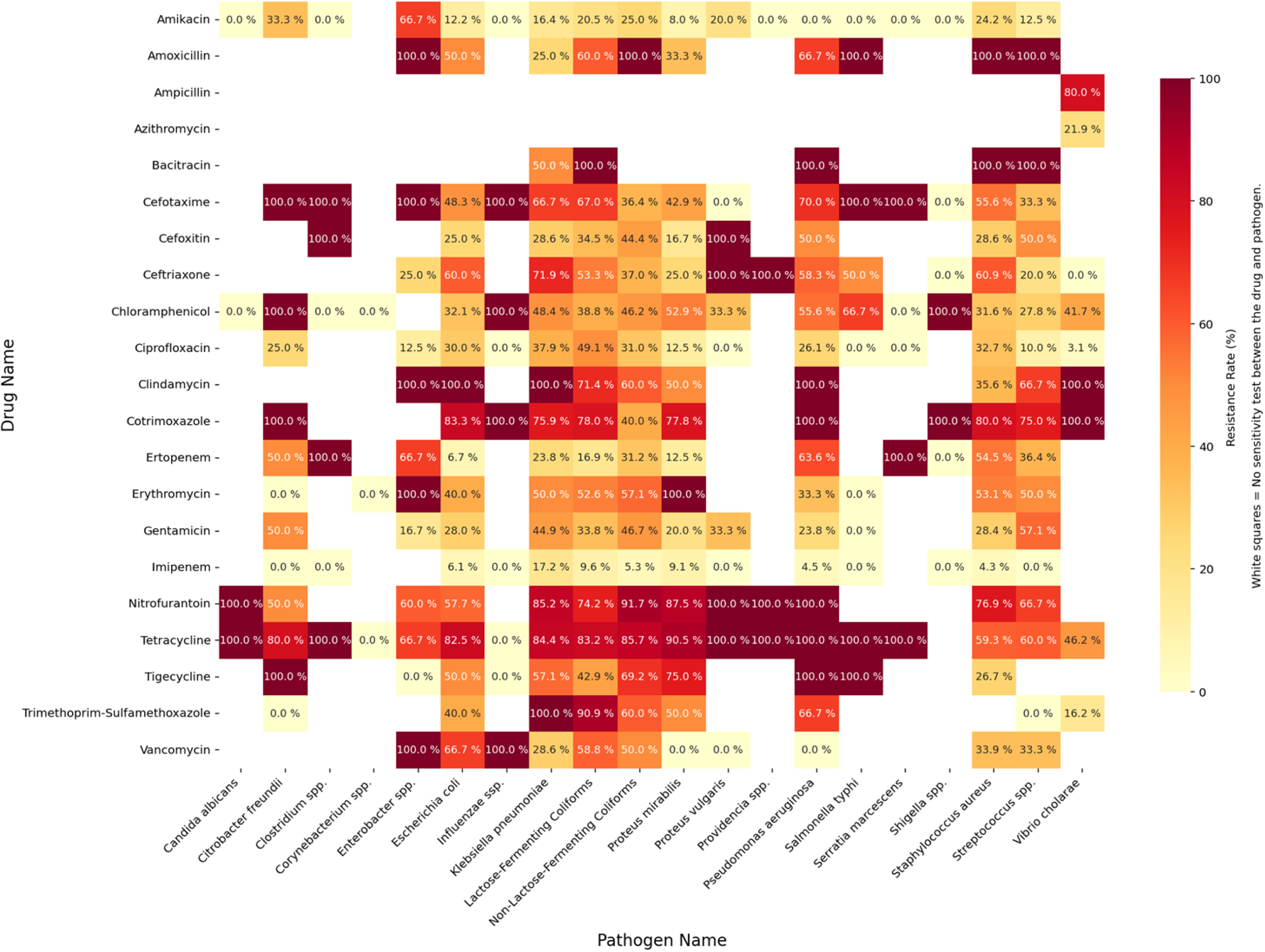
Resistance patterns of antibiotics across pathogens.

MDR prevalence (Figure 11) was highest among *Streptococcus* spp. followed by *K. pneumoniae* (about 500 cases each), whereas *Providencia* spp. exhibited the fewest MDR cases at about 50. The distribution of multidrug-resistant cases reveals that *S. aureus* accounted for the highest burden, followed closely *by Klebsiella* spp. These two pathogens were the dominant contributors to antimicrobial resistance in the dataset. Other notable contributors included *E. coli*, *Pseudomonas* spp., indicating that both Gram-negative and Gram-positive bacteria were significantly involved in the resistance patterns. In contrast, pathogens such as *Shigella* spp., *Salmonella* spp., and *Enterococcus* spp. showed notably fewer cases of resistance.

**Figure 11:**
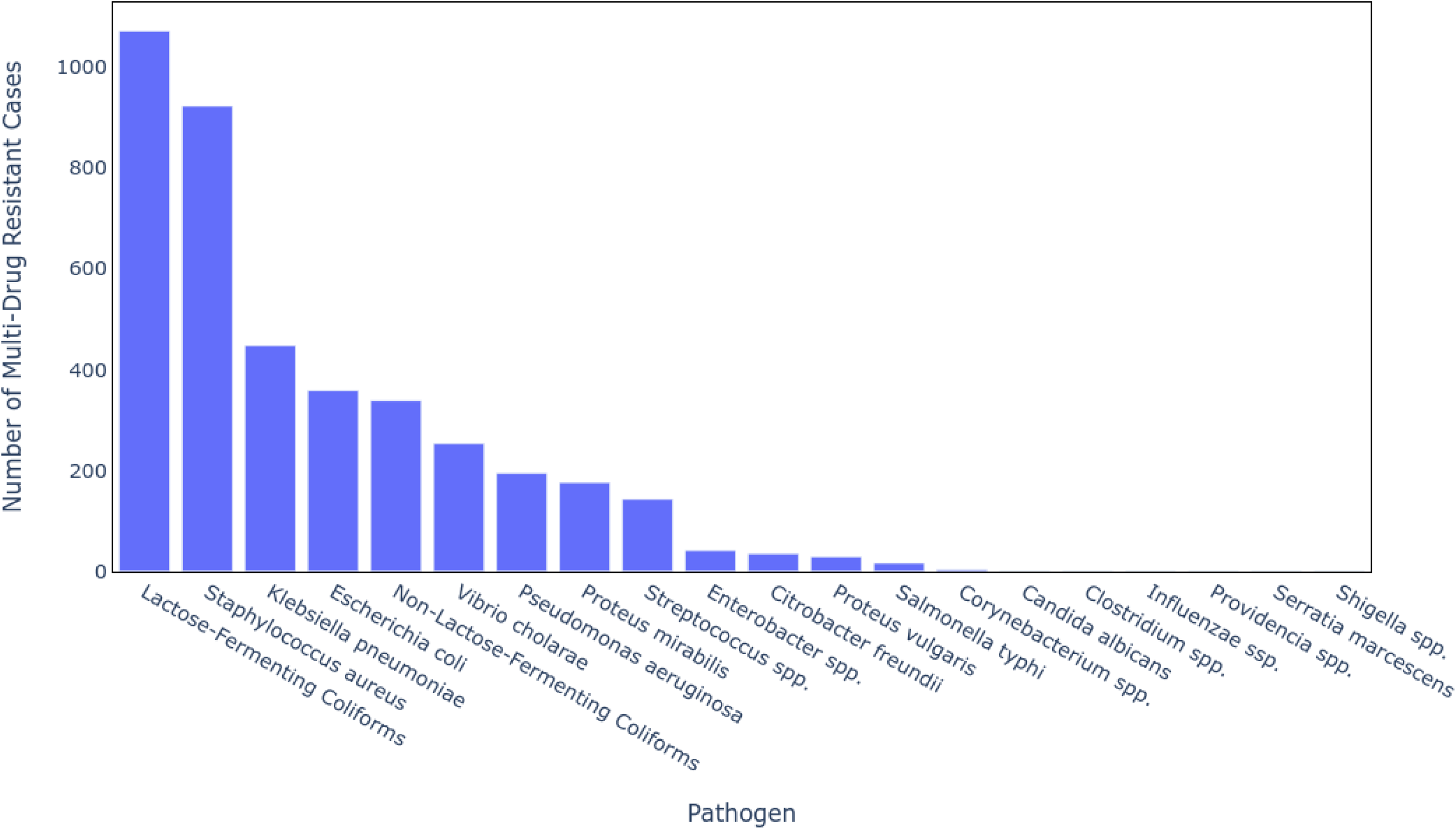
Multidrug resistance counts of various pathogens.

## 4. Discussion

The resistance rates recorded in this study, at 45.5% for males and 41.1% for females, are consistent with international trends of higher resistance in males (Salehi *et al*., 2025). This may be attributed to increased exposure to healthcare facilities or specific risk factors, including catheter use (Salloum *et al*., 2020). The findings of this study are supported by a study conducted in Ethiopia, where similar gender-based disparities were observed, with resistance levels of 48.2% in males and 42.5% in females. This is largely attributed to a higher hospitalisation rate among the male population (Intahphuak *et al*., 2020). The differences could be reflective of general health-seeking behaviours, where males are more often exposed to invasive interventions or antibiotic regimens (Emgård *et al*., 2021). Notably, although our findings align with those of some previous studies, there has been research that reported conflicting results, suggesting that gender may not necessarily affect resistance to certain infections, such as uropathogenic infections (Sié *et al*., 2019).

The results also indicate resistant patterns across age groups, ranging from 35.1% in children (0–4 years) to 45.5% in older individuals (≥65 years), which aligns with reports of older patients having up to 50% resistance rates in the South African population (Chaw *et al*., 2018). This trend can be attributed to comorbidities and cumulative lifetime exposure to antibiotics (Pringle *et al*., 2017), and it calls for tailored antibiotic stewardship strategies that target age-specific risk factors (Bansal *et al*., 2019). Additionally, Kosai *et al*. (2017) confirm that the prevalence of antibiotic resistance increases with age. In contrast, there is evidence from some previous studies indicating that antibiotic resistance in younger patients could be worse due to their higher exposure to antibiotics in the earlier phases of life (Wu *et al*., 2021).

Our results showed significant variations among seasons in resistance rates, with 45.4% resistance in winter and 41.6% in summer, aligning with the findings of previous studies that suggest higher resistance rates in colder months (Fischer and Bild, 2019). The influence of seasonal dynamics on infection indicates a complex interplay between environmental dynamics and resistance levels. Therefore, high vigilance during winter seasons is paramount (Shapiro *et al*., 2019). Additionally, this requires the development of integrated surveillance systems that can detect and respond to seasonal trends in resistance (Abeleda *et al*., 2023).

The extreme differences in resistance across wards, increasing to 75% in stroke wards from just 25% in postnatal wards, are a cause for concern. The differences could be due to the severity of illness in patients and the level of exposure to antibiotics across various clinical settings (Zahedani *et al*., 2018). Previous studies have also reported that specialty wards and ICUs have higher levels of resistance, likely due to longer hospital stays and more extensive use of antibiotics (Wu and Zhao, 2023). These findings call for targeted intervention in high-risk environments, such as improved infection control practices in stroke wards and ICU environments (Sholeh *et al*., 2021).

Our results indicate alarming antimicrobial resistance patterns recorded in this study, for example, 100% resistance of *P. mirabilis* and intermediate susceptibility of *K. pneumoniae* (75% were susceptible). Comparable AMR levels have been reported in Kenya, with up to 95% resistance of *P. mirabilis* to multiple common antibiotics, highlighting the global threat posed by this pathogen, often augmented by plasmid-mediated resistance genes (Fischer and Bild, 2019; Van Bijnen *et al*., 2015). Additionally, class-level trends for antibiotic effectiveness, characterised by 90% effectiveness of imipenem vs 10% effectiveness of Bactrim, echo worldwide reports of Bactrim resistance exceeding 70% in most areas (Bryce *et al*., 2018). These findings underscore the urgent need to update treatment guidelines based on real-time data regarding resistance patterns (Goldstein *et al*., 2018).

Analysis of the distribution of MDR cases revealed that *S. aureus* and *K. pneumoniae* accounted for the highest burden of resistance, emerging as the dominant contributors to the AMR profile. These were followed by other notable pathogens, such as *E. coli* and *Pseudomonas* spp., highlighting the importance of both Gram-positive and Gram-negative bacteria in influencing resistance patterns. In contrast, fewer MDR cases were recorded among *Shigella* spp., *Salmonella* spp., and *Enterococcus* spp., suggesting a much lower resistance burden within these groups. The concentration of resistance in a few high-priority organisms signals the need for focused surveillance and intervention strategies.

These findings corroborate those of previous studies in Zimbabwe, such as Mhondoro *et al*. (2019), which reported high resistance rates in *E. coli* and *S. aureus*, and high resistance to last-resort antibiotics, such as fluoroquinolones and carbapenems, in *Salmonella* spp., *P*. *aeruginosa*, and *A. baumannii*. These trends emphasize the pressing necessity of implementing comprehensive AMR surveillance systems to support timely treatment decisions and strengthen antimicrobial stewardship efforts, particularly in resource-limited settings.

Resistance patterns in this study revealed that resistance rates were highest for bacitracin (91.7%), ampicillin (80%), and nitrofurantoin (75.7%), while resistance was moderate for tetracycline (72.1%), amoxicillin (63.8%), and cefotaxime (57.7%). In contrast, resistance to amikacin (17.5%) and imipenem (7.7%) was low, suggesting their continued effectiveness. These trends align with previous findings from Zimbabwe, where high resistance was reported against widely used antibiotics such as ampicillin, penicillin, and ciprofloxacin (Magwenzi *et al*., 2017; WHO, 2018; Llor and Bjerrum, 2015). The 2007 AMR situational analysis identified these drugs among the most prescribed, with resistance increasing between 2012 and 2017 due to overuse (WHO, 2018; Llor and Bjerrum, 2015). The rising resistance to augmentin and cephalosporins might reflect increased reliance on these drugs as alternatives, a trend also reported in South Korea, where third-generation cephalosporin resistance was linked to ESBL-producing bacteria (Kim *et al*., 2017).

Our findings confirm a notable presence of ESBL-producing Enterobacteriaceae, consistent with the findings of Magwenzi *et al*. (2017), who reported 52% ESBL carriage in Harare. Such organisms hydrolyze beta-lactam antibiotics, reducing treatment efficacy (Peirano and Pitout, 2019). Consequently, treatments often require costly fourth-generation cephalosporins and aminoglycosides (Ventola, 2015). Fortunately, Carbapenem resistance remains low (<1%) as indicated in this study and previously in Harare (Magwenzi *et al*., 2017), affirming their status as effective last-line agents despite limited accessibility in low-income settings.

The accurate prediction of resistance patterns enhances the strength of AMR surveillance. The better performance of the SVM model with an accuracy of 72.08% supports previous findings in favour of SVM efficiency in predicting antimicrobial resistance (Bryce *et al*., 2018). In contrast, gradient boosting’s high recall (0.880) and low specificity (0.433) suggest that sensitivity must be traded off against the risks of potential overpredictions in clinical practice (Binsted and McNally, 2024). These findings are in agreement with current research pointing out issues of class imbalance in machine learning algorithms applied in clinical environments (Wu *et al*., 2021).

Our findings have significant implications for AMR surveillance in Gweru and similar resource-limited environments as they validate CDC recommendations for targeted interventions (Alrebish *et al*., 2022). They could inform on the possibility of using ML models, such as SVM, in active surveillance systems, iteratively adjusting empirical therapies as successfully practiced in UK hospitals (Khalid *et al*., 2023). The striking ineffectiveness of currently used antibiotics, such as Bactrim, indicates the need for updated clinical guidelines, resonating with the WHO global AMR action plan.

## 5. Limitations and future work

This study had limitations related to its retrospective secondary analysis study design. The dataset was obtained from routinely collected laboratory reports; records of key clinical information and distinction between hospital-acquired and community-acquired infections were not available. It was also impossible to ascertain whether the reported AMR was primary or secondary. Therefore, resistance trends were therefore used over the entire study population without stratification by acquisition site or patient visit history. In addition, while data were collected at a public sector tertiary hospital, they are not inherently representative of AMR patterns at nearby rural clinics or private health facilities, which could compromise the generalizability of the results to other health facilities within the Midlands Province or Zimbabwe at large.

Further research should involve expanding AMR surveillance efforts within Zimbabwean hospitals using linked networks to generate comprehensive datasets necessary to maximize ML model efficiency. The incorporation of genetic sequencing to characterize resistance mechanisms will be critical in maximizing predictive models, supported by affordable technologies. Comprehensive longitudinal research is required to track evolving AMR patterns and inform adaptive treatment policies.

## 6. Conclusion

This study, based on clinical and demographic data from 874 patients, demonstrated the potential of machine learning to enhance antimicrobial resistance surveillance in Gweru, Zimbabwe. The SVM model was developed and achieved an accuracy of 72.08%, precision of 73.25%, and recall rate of 79.78%, outperforming the random forest, logistic regression, gradient boosting, and KNN models. Antibiotic use was identified as the primary driver of AMR, with hospital ward, patient age, and pathogen type as significant factors, revealing high MDR in *S. aureus* and low MDR in *Shigella* spp. and *Serratia marcescens*. These findings highlight the value of machine learning-driven tools for improving AMR surveillance and stewardship in resource-limited settings, where traditional surveillance methods are weak. This framework can inform national AMR strategies by enabling low-cost and scalable surveillance solutions.

### Ethical approval

Ethical approval for this study was granted by the institutional review boards and ethics committees of the Gweru Provincial Hospital. The patient records that formed the main dataset were completely anonymized so that the participant identities were not recognizable. The board waived the requirement for informed consent because of the retrospective nature of the study. Data were kept confidential, securely stored and only used for the purpose of this study. Administrative approval to access the data was granted by the same body.

## Authors’ contributions

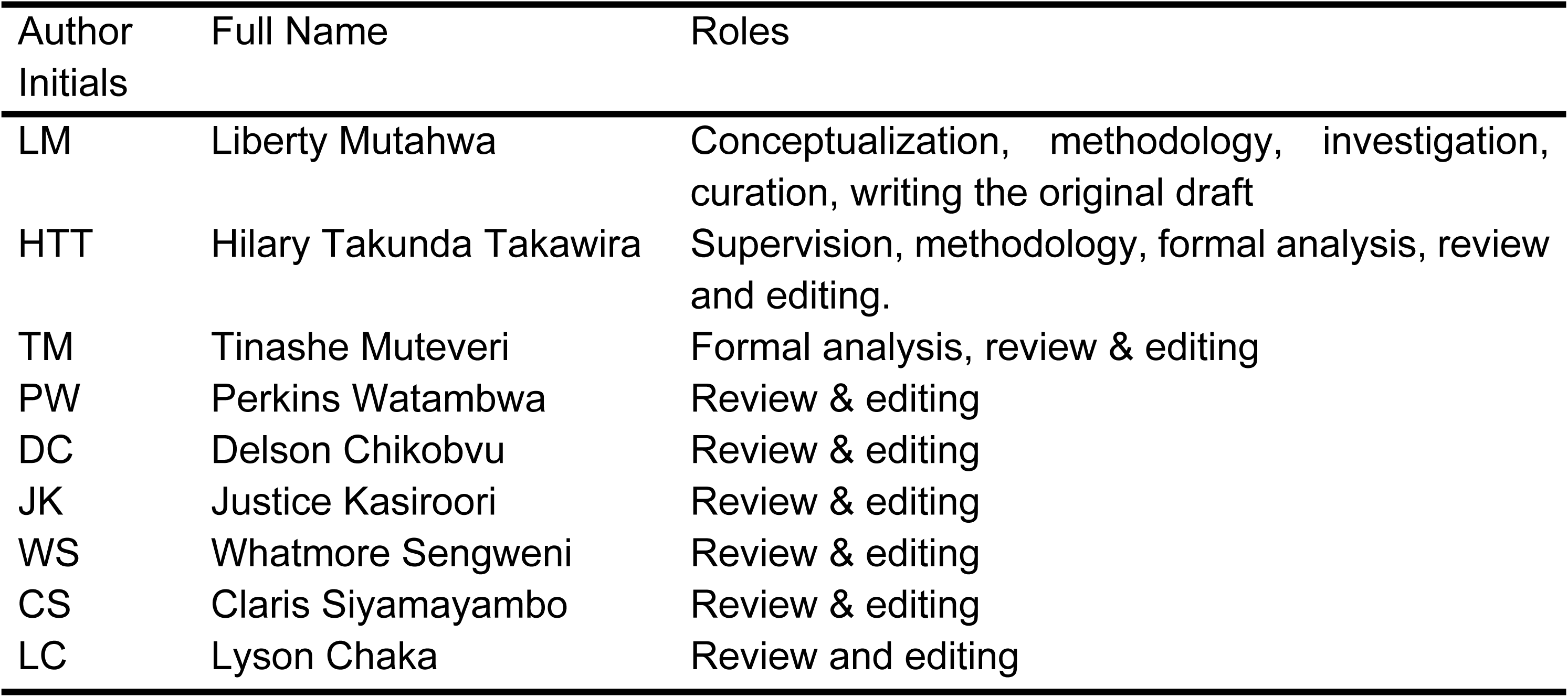

## Data availability

Due to patient confidentiality and ethical restrictions, the dataset is not publicly available. De-identified data may be accessed upon reasonable request.

## References

Abeleda, M. A., Peña, I., Salenga, R., Capule, F., Nacabu-an, S. M. and Nala, P. (2023). ‘Antimicrobial Consumption and Resistance of Restricted Antibiotics in a Level III Government Hospital’. Acta Medica Philippina. doi: 10.47895/amp.vi0.8056.

Adedeji, R., Agboeze, T., Akomolafe, A. and Daramola, O. (2025). ‘Supervised Learning Model Systems to Predict and Identify Drivers of AMR in Africa’. Wellcome Open Research, 10, p. 410. doi: 10.12688/wellcomeopenres.24135.1.

Africa CDC (2023) Antimicrobial resistance is a greater threat than HIV/AIDS, TB, and malaria, says new report. Available at: https://africacdc.org/news-item/antimicrobial-resistance-is-a-greater-threat-than-hiv-aids-tb-and-malaria-says-new-report/

Ali, T., Ahmed, S. and Aslam, M. (2023). “Artificial Intelligence for Antimicrobial Resistance Prediction: Challenges and Opportunities towards Practical Implementation.” Antibiotics, 12 (3), p. 523. doi: 10.3390/antibiotics12030523

Alrebish, S. A., Ahmed, N. J., Al Hamed, H., Kumar, A., Yusufoglu, H. S. and Khan, A. H. (2022). ‘Antibiotic Susceptibility of Bacterial Pathogens Stratified by Age in a Public Hospital in Qassim’. Healthcare, 10 (9), p. 1757. doi: 10.3390/healthcare10091757.

Aruhomukama, D. and Nakabuye, H. (2023). ‘Investigating the evolution and predicting the future outlook of antimicrobial resistance in sub-saharan Africa using phenotypic data for Klebsiella pneumoniae: a 12-year analysis’. BMC Microbiology, 23 (1), p. 214. doi: 10.1186/s12866-023-02966-y.

Asadi, G., Aslani, A., Nayebinia, A.-S. and Fathnezhad-Kazemi, A. (2020). ‘Explaining breastfeeding experiences and assessing factors affecting breastfeeding self-efficacy in mothers of premature infants: a mixed method study protocol’. Reproductive Health, 17 (1), p. 42. doi: 10.1186/s12978-020-0895-2.

Aslam, B., Asghar, R., Muzammil, S., Shafique, M., Siddique, A. B., Khurshid, M., Ijaz, M., Rasool, M. H., Chaudhry, T. H., Aamir, A. and Baloch, Z. (2024). ‘AMR and Sustainable Development Goals: at a crossroads’. Globalization and Health, 20 (1), p. 73. doi: 10.1186/s12992-024-01046-8.

Babirye, S. R., Nsubuga, M., Mboowa, G., Batte, C., Galiwango, R. and Kateete, D. P. (2024). ‘Machine learning-based prediction of antibiotic resistance in Mycobacterium tuberculosis clinical isolates from Uganda’. BMC Infectious Diseases, 24 (1), p. 1391. doi: 10.1186/s12879-024-10282-7.

Bansal, R., Jain, A., Goyal, M., Singh, T., Sood, H. and Malviya, H. (2019). ‘Antibiotic abuse during endodontic treatment: A contributing factor to antibiotic resistance’. Journal of Family Medicine and Primary Care, 8 (11), p. 3518. doi: 10.4103/jfmpc.jfmpc_768_19.

Benkwitz-Bedford, S., Palm, M., Demirtas, T. Y., Mustonen, V., Farewell, A., Warringer, J., Parts, L. and Moradigaravand, D. (2021). ‘Machine Learning Prediction of Resistance to Subinhibitory Antimicrobial Concentrations from Escherichia coli Genomes’. mSystems. Edited by T. A. Van Laar, 6 (4), p. 10.1128/msystems.00346-21. doi: 10.1128/msystems.00346-21.

Binsted, L. E. and McNally, L. (2024). ‘Diverse Relationships Between Antibiotic Resistance and Host Age: A Meta-Analysis Across Antibiotic Classes and Bacterial Genera’. Epidemiology. doi: 10.1101/2024.02.25.24303263.

Bo, L., Sun, H., Li, Y.-D., Zhu, J., Wurpel, J. N. D., Lin, H. and Chen, Z.-S. (2024). ‘Combating antimicrobial resistance: the silent war’. Frontiers in Pharmacology, 15, p. 1347750. doi: 10.3389/fphar.2024.1347750.

Broschat, S. L., Siu, S. W. I. and De La Fuente-Nunez, C. (2024). ‘Editorial: Machine learning approaches to antimicrobials: discovery and resistance’. Frontiers in Bioinformatics, 4, p. 1458237. doi: 10.3389/fbinf.2024.1458237.

Bryce, A., Costelloe, C., Wootton, M., Butler, C. C. and Hay, A. D. (2018). ‘Comparison of risk factors for, and prevalence of, antibiotic resistance in contaminating and pathogenic urinary Escherichia coli in children in primary care: prospective cohort study’. Journal of Antimicrobial Chemotherapy, 73 (5), pp. 1359–1367. doi: 10.1093/jac/dkx525.

Camacho, D. M., Collins, K. M., Powers, R. K., Costello, J. C. and Collins, J. J. (2018). ‘Next-Generation Machine Learning for Biological Networks’. Cell, 173 (7), pp. 1581– 1592. doi: 10.1016/j.cell.2018.05.015.

Chaw, P. S., Schlinkmann, K. M., Raupach-Rosin, H., Karch, A., Pletz, M. W., Huebner, J., Nyan, O. and Mikolajczyk, R. (2018). ‘Antibiotic use on paediatric inpatients in a teaching hospital in the Gambia, a retrospective study’. Antimicrobial Resistance & Infection Control, 7 (1), p. 82. doi: 10.1186/s13756-018-0380-7.

Chetty, S., Reddy, M., Ramsamy, Y., Dlamini, V. C., Reddy-Naidoo, R. and Essack, S. Y. (2022). ‘Antimicrobial Stewardship in Public-Sector Hospitals in KwaZulu-Natal, South Africa’. Antibiotics, 11 (7), p. 881. doi: 10.3390/antibiotics11070881.

Chiwodza, Z., Varney, J., Jr, Zvinoera, K. and Ndhlovu, K. (2023). ‘AMR Trends in UTI: The need for Laboratory, Pharmacy and Clinic Collaboration at a Provincial Hospital’. The Global Health Network Collections. doi: 10.21428/3d48c34a.34a05bef.

Emgård, M., Mwangi, R., Mayo, C., Mshana, E., Nkini, G., Andersson, R., Msuya, S. E., Lepp, M., Muro, F. and Skovbjerg, S. (2021). ‘Tanzanian primary healthcare workers’ experiences of antibiotic prescription and understanding of antibiotic resistance in common childhood infections: a qualitative phenomenographic study’. Antimicrobial Resistance & Infection Control, 10 (1), p. 94. doi: 10.1186/s13756-021-00952-5.

Fadare, J. O., Ogunleye, O., Iliyasu, G., Adeoti, A., Schellack, N., Engler, D., Massele, A. and Godman, B. (2019). ‘Status of antimicrobial stewardship programmes in Nigerian tertiary healthcare facilities: Findings and implications’. Journal of Global Antimicrobial Resistance, 17, pp. 132–136. doi: 10.1016/j.jgar.2018.11.025.

Feretzakis, G., Loupelis, E., Sakagianni, A., Kalles, D., Martsoukou, M., Lada, M., Skarmoutsou, N., Christopoulos, C., Valakis, K., Velentza, A., Petropoulou, S., Michelidou, S. and Alexiou, K. (2020). ‘Using Machine Learning Techniques to Aid Empirical Antibiotic Therapy Decisions in the Intensive Care Unit of a General Hospital in Greece’. Antibiotics, 9 (2), p. 50. doi: 10.3390/antibiotics9020050.

Fischer, M. M. and Bild, M. (2019). ‘Hospital use of antibiotics as the main driver of infections with antibiotic-resistant bacteria – a reanalysis of recent data from the European Union’. Microbiology. doi: 10.1101/553537.

Ganaie, M. A., Hu, M., Malik, A. K., Tanveer, M. and Suganthan, P. N. (2022). ‘Ensemble deep learning: A review’. Engineering Applications of Artificial Intelligence, 115, p. 105151. doi: 10.1016/j.engappai.2022.105151.

Goldstein, E., MacFadden, D. R., Lee, R. S. and Lipsitch, M. (2018). ‘Outpatient prescribing of four major antibiotic classes and prevalence of antimicrobial resistance in US adults’. Epidemiology. doi: 10.1101/456244.

Gulumbe, B. H., Haruna, U. A., Almazan, J., Ibrahim, I. H., Faggo, A. A. and Bazata, A. Y. (2022). ‘Combating the menace of antimicrobial resistance in Africa: a review on stewardship, surveillance and diagnostic strategies’. Biological Procedures Online, 24 (1), p. 19. doi: 10.1186/s12575-022-00182-y.

Intahphuak, S., Apidechkul, T. and Kuipiaphum, P. (2020). ‘Antibiotic resistance among the Lahu hill tribe people in northern Thailand: a cross-sectional study’. In Review. doi: 10.21203/rs.3.rs-27927/v1.

Institute for Health Metrics and Evaluation (2023) Zimbabwe. Available at: https://www.healthdata.org/sites/default/files/2023-09/Zimbabwe.pdf

Iskandar, K., Molinier, L., Hallit, S., Sartelli, M., Hardcastle, T. C., Haque, M., Lugova, H., Dhingra, S., Sharma, P., Islam, S., Mohammed, I., Naina Mohamed, I., Hanna, P. A., Hajj, S. E., Jamaluddin, N. A. H., Salameh, P. and Roques, C. (2021). ‘Surveillance of antimicrobial resistance in low- and middle-income countries: a scattered picture’. Antimicrobial Resistance & Infection Control, 10 (1), p. 63. doi: 10.1186/s13756-021-00931-w.

Kariuki, S., Kering, K., Wairimu, C., Onsare, R. and Mbae, C. (2022). ‘Antimicrobial Resistance Rates and Surveillance in Sub-Saharan Africa: Where Are We Now?’ Infection and Drug Resistance, Volume 15, pp. 3589–3609. doi: 10.2147/IDR.S342753.

Kasew, D., Eshetie, S., Diress, A., Tegegne, Z. and Moges, F. (2021). ‘Multiple drug resistance bacterial isolates and associated factors among urinary stone patients at the University of Gondar Comprehensive Specialized Hospital, Northwest Ethiopia’. BMC Urology, 21 (1), p. 27. doi: 10.1186/s12894-021-00794-8.

Khalid, N., Qayyum, A., Bilal, M., Al-Fuqaha, A. and Qadir, J. (2023). ‘Privacy-preserving artificial intelligence in healthcare: Techniques and applications’. Computers in Biology and Medicine, 158, p. 106848. doi: 10.1016/j.compbiomed.2023.106848.

Kim, D., Ahn, J. Y., Lee, C. H., Jang, S. J., Lee, H., Yong, D., Jeong, S. H. and Lee, K. (2017). ‘Increasing Resistance to Extended-Spectrum Cephalosporins, Fluoroquinolone, and Carbapenem in Gram-Negative Bacilli and the Emergence of Carbapenem Non-Susceptibility in Klebsiella pneumoniae : Analysis of Korean Antimicrobial Resistance Monitoring System (KARMS) Data From 2013 to 2015’. Annals of Laboratory Medicine, 37 (3), pp. 231–239. doi: 10.3343/alm.2017.37.3.231.

Kim, J. I., Maguire, F., Tsang, K. K., Gouliouris, T., Peacock, S. J., McAllister, T. A., McArthur, A. G. and Beiko, R. G. (2022). ‘Machine Learning for Antimicrobial Resistance Prediction: Current Practice, Limitations, and Clinical Perspective’. Clinical Microbiology Reviews, 35 (3), pp. e00179–21. doi: 10.1128/cmr.00179-21.

Kolluru, V. K., Nuthakki, Y., Koganti, S. and Chintakunta, A. N. (2024). ‘Use of Predictive Analytics in Antimicrobial Resistance: A Review’. Cognizance Journal of Multidisciplinary Studies, 4 (1), pp. 404–414. doi: 10.47760/cognizance.2024.v04i01.020.

Kosai, K., Kaku, N., Uno, N., Saijo, T., Morinaga, Y., Imamura, Y., Hasegawa, H., Miyazaki, T., Izumikawa, K., Mukae, H. and Yanagihara, K. (2017). ‘Risk Factors for Acquisition of Fluoroquinolone or Aminoglycoside Resistance in Addition to Carbapenem Resistance in Pseudomonas aeruginosa’. The Open Microbiology Journal, 11 (1), pp. 92–97. doi: 10.2174/1874285801711010092.

Llor, C. and Bjerrum, L. (2014). ‘Antimicrobial resistance: risk associated with antibiotic overuse and initiatives to reduce the problem’. Therapeutic Advances in Drug Safety, 5 (6), pp. 229–241. doi: 10.1177/2042098614554919.

Magwenzi, M. T., Gudza-Mugabe, M., Mujuru, H. A., Dangarembizi-Bwakura, M., Robertson, V. and Aiken, A. M. (2017). ‘Carriage of antibiotic-resistant Enterobacteriaceae in hospitalised children in tertiary hospitals in Harare, Zimbabwe’. Antimicrobial Resistance & Infection Control, 6 (1), p. 10. doi: 10.1186/s13756-016-0155-y.

Mhondoro, M., Ndlovu, N., Bangure, D., Juru, T., Gombe, N. T., Shambira, G., Nsubuga, P. and Tshimanga, M. (2019). ‘Trends in antimicrobial resistance of bacterial pathogens in Harare, Zimbabwe, 2012–2017: a secondary dataset analysis’. BMC Infectious Diseases, 19 (1), p. 746. doi: 10.1186/s12879-019-4295-6.

Murray, C. J. L., Ikuta, K. S., Sharara, F., Swetschinski, L., Robles Aguilar, G., Gray, A., Han, C., Bisignano, C., Rao, P., Wool, E., Johnson, S. C., Browne, A. J., Chipeta, M. G., Fell, F., Hackett, S., Haines-Woodhouse, G., Kashef Hamadani, B. H., Kumaran, E. A. P., McManigal, B., Achalapong, S., Agarwal, R., Akech, S., Albertson, S., Amuasi, J., Andrews, J., Aravkin, A., Ashley, E., Babin, F.-X., Bailey, F., Baker, S., Basnyat, B., Bekker, A., Bender, R., Berkley, J. A., Bethou, A., Bielicki, J., Boonkasidecha, S., Bukosia, J., Carvalheiro, C., Castañeda-Orjuela, C., Chansamouth, V., Chaurasia, S., Chiurchiù, S., Chowdhury, F., Clotaire Donatien, R., Cook, A. J., Cooper, B., Cressey, T. R., Criollo-Mora, E., Cunningham, M., Darboe, S., Day, N. P. J., De Luca, M., Dokova, K., Dramowski, A., Dunachie, S. J., Duong Bich, T., Eckmanns, T., Eibach, D., Emami, A., Feasey, N., Fisher-Pearson, N., Forrest, K., Garcia, C., Garrett, D., Gastmeier, P., Giref, A. Z., Greer, R. C., Gupta, V., Haller, S., Haselbeck, A., Hay, S. I., Holm, M., Hopkins, S., Hsia, Y., Iregbu, K. C., Jacobs, J., Jarovsky, D., Javanmardi, F., Jenney, A. W. J., Khorana, M., Khusuwan, S., Kissoon, N., Kobeissi, E., Kostyanev, T., Krapp, F., Krumkamp, R., Kumar, A., Kyu, H. H., Lim, C., Lim, K., Limmathurotsakul, D., Loftus, M. J., Lunn, M., Ma, J., Manoharan, A., Marks, F., May, J., Mayxay, M., Mturi, N., Munera-Huertas, T., Musicha, P., Musila, L. A., Mussi-Pinhata, M. M., Naidu, R. N., Nakamura, T., Nanavati, R., Nangia, S., Newton, P., Ngoun, C., Novotney, A., Nwakanma, D., Obiero, C. W., Ochoa, T. J., Olivas-Martinez, A., Olliaro, P., Ooko, E., Ortiz-Brizuela, E., Ounchanum, P., Pak, G. D., Paredes, J. L., Peleg, A. Y., Perrone, C., Phe, T., Phommasone, K., Plakkal, N., Ponce-de-Leon, A., Raad, M., Ramdin, T., Rattanavong, S., Riddell, A., Roberts, T., Robotham, J. V., Roca, A., Rosenthal, V. D., Rudd, K. E., Russell, N., Sader, H. S., Saengchan, W., Schnall, J., Scott, J. A. G., Seekaew, S., Sharland, M., Shivamallappa, M., Sifuentes-Osornio, J., Simpson, A. J., Steenkeste, N., Stewardson, A. J., Stoeva, T., Tasak, N., Thaiprakong, A., Thwaites, G., Tigoi, C., Turner, C., Turner, P., Van Doorn, H. R., Velaphi, S., Vongpradith, A., Vongsouvath, M., Vu, H., Walsh, T., Walson, J. L., Waner, S., Wangrangsimakul, T., Wannapinij, P., Wozniak, T., Young Sharma, T. E. M. W., Yu, K. C., Zheng, P., Sartorius, B., Lopez, A. D., Stergachis, A., Moore, C., Dolecek, C. and Naghavi, M. (2022). ‘Global burden of bacterial antimicrobial resistance in 2019: a systematic analysis’. The Lancet, 399 (10325), pp. 629–655. doi: 10.1016/S0140-6736(21)02724-0.

Nirmala, B., Singh, V. and Omar, B. J. (2025). ‘Seasonal Trends and Antibiotic Resistance Profiles of Bacterial Pathogens in Indian Clinical Isolates’. Cureus. doi: 10.7759/cureus.77255.

Nsubuga, M., Galiwango, R., Jjingo, D. and Mboowa, G. (2024). ‘Generalizability of machine learning in predicting antimicrobial resistance in E. coli: a multi-country case study in Africa’. BMC Genomics, 25 (1), p. 287. doi: 10.1186/s12864-024-10214-4.

Okolie, O. J., Ismail, S. U., Igwe, U. and Adukwu, E. C. (2025). ‘Assessing barriers and opportunities for the improvement of laboratory performance and robust surveillance of antimicrobial resistance in Nigeria– a quantitative study’. Antimicrobial Resistance & Infection Control, 14 (1), p. 29. doi: 10.1186/s13756-025-01530-9.

Pearcy, N., Hu, Y., Baker, M., Maciel-Guerra, A., Xue, N., Wang, W., Kaler, J., Peng, Z., Li, F. and Dottorini, T. (2021). ‘Genome-Scale Metabolic Models and Machine Learning Reveal Genetic Determinants of Antibiotic Resistance in Escherichia coli and Unravel the Underlying Metabolic Adaptation Mechanisms’. mSystems. Edited by X. Lin, 6 (4), p. 10.1128/msystems.00913-20. doi: 10.1128/msystems.00913-20.

Peirano, G. and Pitout, J. D. D. (2019). “Extended-Spectrum Β-Lactamase-Producing enterobacteriaceae: Update on molecular epidemiology and treatment options.” Drugs, 79 (14), pp. 1529–1541. doi: 10.1007/s40265-019-01180-3.

Pringle, S., Simpson, M., Nielsen, S., Cooper, C. and Vanniasinkam, T. (2017). ‘Antibiotic prescribing practices in aged care facilities in regional NSW and the ACT’. Journal of Pharmacy Practice and Research, 47 (5), pp. 365–374. doi: 10.1002/jppr.1262.

Quoc, V. T., Ngoc, D. N. T., Hoang, T. N., Thi, H. V., Duc, M. T., Nguyet, T. D. P., Van, T. N., Ngoc, D. H., Son, G. V. and Duc, T. B. (2023). “Predicting antibiotic resistance in ICUs patients by applying machine learning in Vietnam.” Infection and Drug Resistance, Volume 16, pp. 5535–5546. doi: 10.2147/IDR.S415885

Ren, Y., Chakraborty, T., Doijad, S., Falgenhauer, L., Falgenhauer, J., Goesmann, A., Schwengers, O. and Heider, D. (2022). ‘Deep Transfer Learning Enables Robust Prediction of Antimicrobial Resistance for Novel Antibiotics’. Antibiotics, 11 (11), p. 1611. doi: 10.3390/antibiotics11111611.

Richet, H. (2012). ‘Seasonality in Gram-negative and healthcare-associated infections’. Clinical Microbiology and Infection, 18 (10), pp. 934–940. doi: 10.1111/j.1469-0691.2012.03954.x.

Sakagianni, A., Koufopoulou, C., Feretzakis, G., Kalles, D., Verykios, V. S., Myrianthefs, P. and Fildisis, G. (2023). ‘Using Machine Learning to Predict Antimicrobial Resistance―A Literature Review’. Antibiotics, 12 (3), p. 452. doi: 10.3390/antibiotics12030452.

Sakagianni, A., Koufopoulou, C., Koufopoulos, P., Kalantzi, S., Theodorakis, N., Nikolaou, M., Paxinou, E., Kalles, D., Verykios, V. S., Myrianthefs, P. and Feretzakis, G. (2024). ‘Data-Driven Approaches in Antimicrobial Resistance: Machine Learning Solutions’. Antibiotics, 13 (11), p. 1052. doi: 10.3390/antibiotics13111052.

Salehi, M., Laitinen, V., Bhanushali, S., Bengtsson-Palme, J., Collignon, P., Beggs, J. J., Pärnänen, K. and Lahti, L. (2025). ‘Gender differences in global antimicrobial resistance’. npj Biofilms and Microbiomes, 11 (1), p. 79. doi: 10.1038/s41522-025-00715-9.

Salloum, S., Tawk, M. and Tayyara, L. (2020). ‘Bacterial resistance to antibiotics and associated factors in two hospital centers in Lebanon from January 2017 to June 2017’. Infection Prevention in Practice, 2 (2), p. 100043. doi: 10.1016/j.infpip.2020.100043.

Sartorius, B., Gray, A. P., Davis Weaver, N., Robles Aguilar, G., Swetschinski, L. R., Ikuta, K. S., Mestrovic, T., Chung, E., Wool, E. E., Han, C., Gershberg Hayoon, A., Araki, D. T., Abd-Elsalam, S., Aboagye, R. G., Adamu, L. H., Adepoju, A. V., Ahmed, A., Akalu, G. T., Akande-Sholabi, W., Amuasi, J. H., Amusa, G. A., Argaw, A. M., Aruleba, R. T., Awoke, T., Ayalew, M. K., Azzam, A. Y., Babin, F.-X., Banerjee, I., Basiru, A., Bayileyegn, N. S., Belete, M. A., Berkley, J. A., Bielicki, J. A., Dekker, D., Demeke, D., Demsie, D. G., Dessie, A. M., Dunachie, S. J., Ed-Dra, A., Ekholuenetale, M., Ekundayo, T. C., El Sayed, I., Elhadi, M., Elsohaby, I., Eyre, D., Fagbamigbe, A. F., Feasey, N. A., Fekadu, G., Fell, F., Forrest, K. M., Gebrehiwot, M., Gezae, K. E., Ghazy, R. M., Hailegiyorgis, T. T., Haines-Woodhouse, G., Hasaballah, A. I., Haselbeck, A. H., Hsia, Y., Iradukunda, A., Iregbu, K. C., Iwu, C. C. D., Iwu-Jaja, C. J., Iyasu, A. N., Jaiteh, F., Jeon, H., Joshua, C. E., Kassa, G. G., Katoto, P. D., Krumkamp, R., Kumaran, E. A. P., Kyu, H. H., Manilal, A., Marks, F., May, J., McLaughlin, S. A., McManigal, B., Melese, A., Misgina, K. H., Mohamed, N. S., Mohammed, M., Mohammed, Shafiu, Mohammed, Shikur, Mokdad, A. H., Moore, C. E., Mougin, V., Mturi, N., Mulugeta, T., Musaigwa, F., Musicha, P., Musila, L. A., Muthupandian, S., Naghavi, P., Negash, H., Nuckchady, D. C., Obiero, C. W., Odetokun, I. A., Ogundijo, O. A., Okidi, L., Okonji, O. C., Olagunju, A. T., Olufadewa, I. I., Pak, G. D., Perovic, O., Pollard, A., Raad, M., Rafaï, C., Ramadan, H., Redwan, E. M. M., Roca, A., Rosenthal, V. D., Saleh, M. A., Samy, A. M., Sharland, M., Shittu, A., Siddig, E. E., Sisay, E. A., Stergachis, A., Tesfamariam, W. B., Tigoi, C., Tincho, M. B., Tiruye, T. Y., Umeokonkwo, C. D., Walsh, T., Walson, J. L., Yusuf, H., Zeru, N. G., Hay, S. I., Dolecek, C., Murray, C. J. L. and Naghavi, M. (2024). ‘The burden of bacterial antimicrobial resistance in the WHO African region in 2019: a cross-country systematic analysis’. The Lancet Global Health, 12 (2), pp. e201–e216. doi: 10.1016/S2214-109X(23)00539-9.

Shahraki Zahedani, S., Department of Microbiology, School of Medicine, Infectious Diseases and Tropical Medicine Research Center, Zahedan University of Medical Sciences, Zahedan, Iran, Sayadzai, N., and Department of microbiology, Faculty of Medicine, Zahedan University of Medical Sciences, Zahedan, Iran. (2018). ‘Frequency and Antibiotic Resistance Pattern of Diarrheagenic Escherichia coli (DEC) Strains Isolated from Children Aged Less Than 10 Years’. Medical Laboratory Journal, 12 (2), pp. 7–12. doi: 10.29252/mlj.12.2.7.

Shapiro, J. T., Leboucher, G., Myard-Dury, A.-F., Girardo, P., Luzatti, A., Mary, M., Sauzon, J.-F., Lafay, B., Dauwalder, O., Laurent, F., Lina, G., Chidiac, C., Couray-Targe, S., Vandenesch, F., Flandrois, J.-P. and Rasigade, J.-P. (2019). ‘Metapopulation ecology links antibiotic resistance, consumption and patient transfers in a network of hospital wards’. Ecology. doi: 10.1101/771790.

Sholeh, M. A., Kuntaman, K. and Hadi, U. (2021). ‘Quantity of Antibiotic Use and Resistance Pattern of Gut Normal Flora Escherichia coli at Intensive Care Unit and Tropic Infection Ward, Dr Soetomo Hospital, Surabaya, Indonesia’. Folia Medica Indonesiana, 56 (3), p. 159. doi: 10.20473/fmi.v56i3.24472.

Sié, A., Coulibaly, B., Adama, S., Ouermi, L., Dah, C., Tapsoba, C., Bärnighausen, T., Kelly, J. D., Doan, T., Lietman, T. M., Keenan, J. D. and Oldenburg, C. E. (2019). ‘Antibiotic Prescription Patterns among Children Younger than 5 Years in Nouna District, Burkina Faso’. The American Journal of Tropical Medicine and Hygiene, 100 (5), pp. 1121–1124. doi: 10.4269/ajtmh.18-0791.

Sunuwar, J. and Azad, R. K. (2021). ‘A machine learning framework to predict antibiotic resistance traits and yet unknown genes underlying resistance to specific antibiotics in bacterial strains’. Briefings in Bioinformatics, 22 (6), p. bbab179. doi: 10.1093/bib/bbab179.

Van Bijnen, E. M. E., Paget, J., De Lange-de Klerk, E. S. M., Den Heijer, C. D. J., Versporten, A., Stobberingh, E. E., Goossens, H., Schellevis, F. G., and collaboration with the APRES Study Team. (2015). ‘Antibiotic Exposure and Other Risk Factors for Antimicrobial Resistance in Nasal Commensal Staphylococcus aureus: An Ecological Study in 8 European Countries’. PLOS ONE. Edited by A. Cloeckaert, 10 (8), p. e0135094. doi: 10.1371/journal.pone.0135094.

Ventola, C.L. (2015) ‘The antibiotic resistance crisis: part 1: causes and threats’, P & T: A Peer-Reviewed Journal for Formulary Management, 40(4), pp. 277–283.

World Health Organization. (2018). Antimicrobial resistance: Key facts. Retrieved from http://www.who.int/news-room/fact-sheets/detail/antimicrobial-resistance

Wu, H. and Zhao, Q. (2023). ‘Analysis of Multi-Drug Resistant Organism Surveillance and Antimicrobial Resistance Early Warning in a Hospital in 2022’. Journal of Clinical and Nursing Research, 7 (3), pp. 60–69. doi: 10.26689/jcnr.v7i3.4885.

Wu, L., Xie, X., Li, Y., Liang, T., Zhong, H., Ma, J., Yang, L., Yang, J., Li, L., Xi, Y., Li, H., Zhang, J., Chen, X., Ding, Y. and Wu, Q. (2021). ‘Metagenomics-Based Analysis of the Age-Related Cumulative Effect of Antibiotic Resistance Genes in Gut Microbiota’. Antibiotics, 10 (8), p. 1006. doi: 10.3390/antibiotics10081006.

Zhang, X., Hu, Y., Cheng, Z. and Archibald, J. M. (2025). ‘AMRLearn: Protocol for a machine learning pipeline for characterization of antimicrobial resistance determinants in microbial genomic data’. STAR Protocols, 6 (2), p. 103733. doi: 10.1016/j.xpro.2025.103733.

